# Polygenic Risk, Psychopathology, and Personalized Functional Brain Network Topography in Adolescence

**DOI:** 10.1101/2024.09.20.24314007

**Authors:** Kevin Y. Sun, J. Eric Schmitt, Tyler M. Moore, Ran Barzilay, Laura Almasy, Laura M. Schultz, Allyson P. Mackey, Eren Kafadar, Zhiqiang Sha, Jakob Seidlitz, Travis T. Mallard, Zaixu Cui, Hongming Li, Yong Fan, Damien A. Fair, Theodore D. Satterthwaite, Arielle S. Keller, Aaron Alexander-Bloch

**Author notes:** Corresponding Authors: Kevin Y. Sun, BA, MD-PhD Candidate 3700 Hamilton Walk, B504, Aaron Alexander-Bloch, MD, PhD 3700 Hamilton Walk, B504. Senior authors (Arielle S. Keller, Aaron Alexander-Bloch).

## Abstract

**Importance:** Functional brain networks are associated with both behavior and genetic factors. To uncover biological mechanisms of psychopathology, it is critical to define how the spatial organization of these networks relates to genetic risk during development.

**Objective:** To determine the relationships among transdiagnostic polygenic risk scores (PRSs), personalized functional brain networks (PFNs), and overall psychopathology (p-factor) during early adolescence.

**Design:** The Adolescent Brain Cognitive Development (ABCD) Study^lll^ is an ongoing longitudinal cohort study of 21 collection sites across the United States. Here, we conduct a cross-sectional analysis of ABCD baseline data, collected 2017-2018.

**Setting:** The ABCD Study**^®^** is a multi-site community-based study.

**Participants:** The sample is largely recruited through school systems. Exclusion criteria included severe sensory, intellectual, medical, or neurological issues that interfere with protocol and scanner contraindications. Split-half subsets were used for cross-validation, matched on age, ethnicity, family structure, handedness, parental education, site, sex, and anesthesia exposure.

**Exposures:** Polygenic risk scores of transdiagnostic genetic factors F1 (PRS-F1) and F2 (PRS-F2) derived from adults in Psychiatric Genomic Consortium and UK Biobanks datasets. PRS-F1 indexes liability for common psychiatric symptoms and disorders related to mood disturbance; PRS-F2 indexes liability for rarer forms of mental illness characterized by mania and psychosis.

**Main Outcomes and Measures:** (1) P-factor derived from bifactor models of youth-and parent-reported mental health assessments. (2) Person-specific functional brain network topography derived from functional magnetic resonance imaging (fMRI) scans.

**Results:** Total participants included 11,873 youths ages 9-10 years old; 5,678 (47.8%) were female, and the mean (SD) age was 9.92 (0.62) years. PFN topography was found to be heritable (*N*=7,459, 57.1% of vertices *h*^2^ *p_FDR_*<0.05, mean *h*^2^=0.35). PRS-F1 was associated with p-factor (*N*=5,815, *r*=0.12, 95% CI [0.09–0.15], p<0.001). Interindividual differences in functional network topography were associated with p-factor (*N*=7,459, mean *r*=0.12), PRS-F1 (*N*=3,982, mean *r*=0.05), and PRS-F2 (*N*=3,982, mean *r*=0.08). Cortical maps of p-factor and PRS-F1 regression coefficients were highly correlated (*r*=0.7, *p*=0.003).

**Conclusions and Relevance:** Polygenic risk for transdiagnostic adulthood psychopathology is associated with both p-factor and heritable PFN topography during early adolescence. These results advance our understanding of the developmental drivers of psychopathology.

**Key Points:** *Question:* What are the relationships among transdiagnostic polygenic risk scores (PRSs), personalized functional brain networks (PFNs), and overall psychopathology (p-factor) during early adolescence?

*Findings:* In this cross-sectional analysis of the Adolescent Brain Cognitive Development (ABCD) Study^lll^ (*N*=11,873, ages 9-10), we found that a PRS of common mood-related psychopathology in adulthood (PRS-F1) was associated with p-factor during early adolescence. Interindividual differences in p-factor, PRS-F1, and PRS-F2 (capturing more severe psychotic disorders in adulthood) were all robustly associated with PFN topography.

*Meaning:* Polygenic risk for transdiagnostic adulthood psychopathology is associated with both p-factor and PFN topography during early adolescence.

## Introduction

Human cerebral cortex is organized into large-scale functional networks that support perceptual, motor, cognitive, and emotional functions^1,2^. Functional connectivity between and within networks has been shown to explain behavioral^3,4^ and psychiatric symptom^5–7^ variability.

Evidence suggests that these networks are heritable^8–10^ and related to gene expression patterns^11,12^; thus, functional network measures may be intermediate phenotypes of psychiatric genetic risk. However, standard fMRI analyses use group atlases, which assume functional networks’ spatial layouts across cortical structure—their functional topography—is consistent across individuals^1,2^. However, recent work establishes extensive inter-individual variation in functional topography, particularly in association cortex^13–17^. This study uses precision functional mapping to capture person-specific functional neuroanatomy and investigate its relationship to transdiagnostic psychiatric genetic risk and symptom burden in early adolescence.

Precision functional mapping has shown that an individual’s personalized functional network (PFN) topography is highly reproducible, stable, and predictive of cortical activation patterns during fMRI tasks^13,18–20^, offering novel opportunities to investigate relationships with inter-individual topographical differences unobtainable through group atlas approaches^21^.

Accordingly, PFN topography in development has been associated with both cognition^17,22^ and p-factor^23^, a measure of overall psychopathology^24,25^. Patterns of PFN topography are known to be heritable in adulthood^26^, but the genetic basis of PFN topography during youth, and its relationship to psychiatric risk, remains unclear.

To characterize the shared genetic architecture among psychiatric symptoms and disorders, a recent multivariate genome-wide association study (GWAS) used genomic structural equation modeling^27^. This approach identified two transdiagnostic genetic factors, F1 and F2, that explain the majority of genetic variation associated with affective and psychotic psychopathology in European ancestry adults. F1 captures common psychopathology broadly related to mood disturbance, while F2 captures rarer forms of serious mental illness characterized by mania and psychosis. However, it remains unknown how the polygenic risk of F1 and F2 are related to overall psychopathology or brain function during early adolescence.

In this study, we leveraged clinical phenotyping, genotyping, functional neuroimaging, and twin pair enrichment included in the Adolescent Brain Cognitive Development (ABCD) Study baseline acquisition (total *N*=11,873, ages 9-10 years)^28–31^ to investigate the genetic underpinnings of overall psychopathology and functional brain network topography. A bifactor model^32^ of mental health items was used to define p-factor, a broad measure of psychopathology with proven reliability, validity, and utility^33–35^. Based on Mallard et al. 2022^27^, we calculated polygenic risk scores of F1 (PRS-F1) and F2 (PRS-F2) in ABCD. As previously^17,22,23,36^, we used non-negative matrix factorization (NMF) to derive PFNs^37^. We hypothesized that polygenic risk for adulthood psychopathology would be associated with both p-factor and PFN topography during early adolescence. Thus, we sought to investigate (1) the heritability of p-factor^34^ and PFN topography^26^; (2) the relationship between p-factor and PRS-F1 and PRS-F2; and (3) associations of PFN topography with p-factor, PRS-F1, and PRS-F2.

## Methods

### Study Overview

This study leverages the baseline sample from the ABCD BIDS Community Collection^28^, which includes *N*=11,873 youths ages 9-10 years old and their caregivers across 22 sites (**eMethods 1**). Caregivers provided informed consent and each ABCD site received Institutional Review Board approval. Using 125 youth and parent-reported mental health items, the p-factor was derived using a bifactor model^38^ (**Fig 1A, eMethods 2, eTable 1, eFig 1**). Polygenic risk scores of latent genetic factors F1 (PRS-F1), indexing liability for common forms of mood disturbance, and F2 (PRS-F2), indexing liability for psychotic disorders (**Fig 1B**), were calculated based on summary statistics from Mallard et al. 2022^27^ (**eMethods 3**). Owing to current GWAS limitations^39,40^, PRSs were only calculated in a European (EUR) ancestry subsample (*N*=5,815) and were adjusted by regressing out the first ten EUR-ancestry principal components^41^.

**Figure 1.**
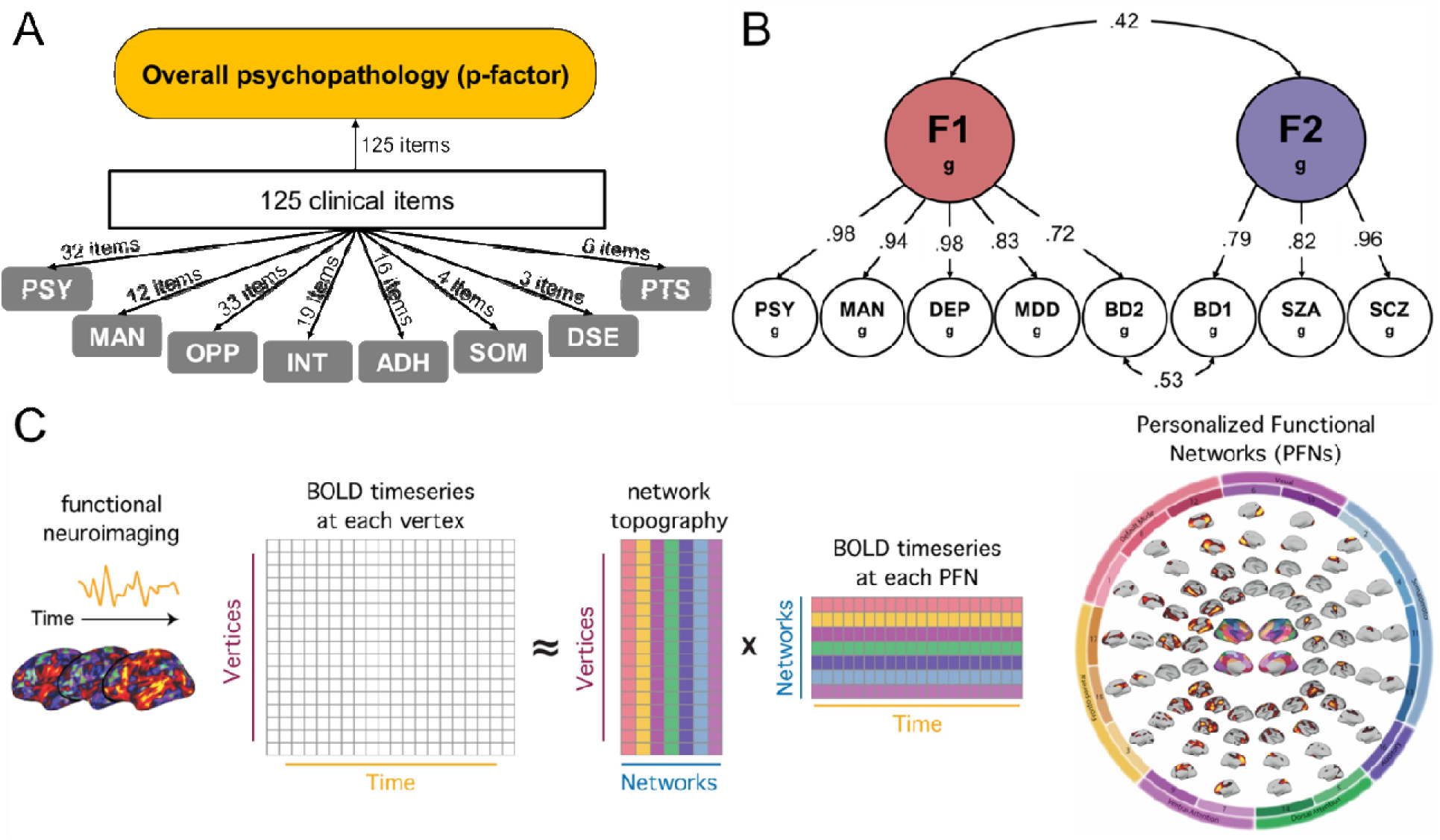
Derivation of P-factor, Polygenic Risk Scores, and Personalized Functional Networks **A.** Overall psychopathology of ABCD participants ages 9-10 was captured using a bifactor model, in which each of 125 mental health interview items loads onto both a general factor (p-factor) and one of 8 orthogonal sub-factors (PSY = psychotic symptoms, MAN = manic symptoms, OPP = oppositional defiance, INT = internalizing symptoms, ADH = attention-deficiency and hyperactivity, SOM = somatic symptoms, DSE = disordered eating, PTS = post-traumatic stress). **B.** In a European ancestry subsample, polygenic risk scores of latent genetic factors F1 (PRS-F1) and F2 (PRS-F2) were calculated based on summary statistics from Mallard et al. 2022^27^ (from which factor structure and loadings are shown here). F1 encapsulates psychiatric symptoms and disorders largely related to mood disturbance (PSY = psychotic symptoms, MAN = manic symptoms, DEP = depressive symptoms, MDD = major depressive disorder, BD2 = bipolar disorder II), and F2 encapsulates rarer, more severe disorders largely related to psychosis (BD1 = bipolar disorder I, SZA = schizoaffective disorder, SCZ = schizophrenia). **C.** Spatially constrained non-negative matrix factorization (NMF) was used to decompose each participant’s fMRI time series into personalized functional networks (PFNs), as defined by a loading matrix of 17 x 59,412 for each participant, in which 17 corresponds to the number of networks and 59,412 corresponds to the number of cortical vertices. Each value in the loading matrix quantifies the extent to which each vertex belongs to a certain network for that participant, a probabilistic network definition we refer to as PFN topography. Schematic reproduced from Keller et al. 2024^36^.

### Personalized Functional Networks (PFNs)

Neuroimaging data was processed using the ABCD-BIDS pipeline^42^ (**eMethods 4**).

Consistent with previous work^22,36^, we concatenated the time series data from up to four resting-state scans and three task-based scans^20^, and excluded participants with incomplete data or excessive head motion, yielding a final sample of *N*=7,459. Mean framewise displacement (FD) was calculated for each participant’s concatenated time series to summarize in-scanner motion and used a model covariate.

PFN generation was consistent with prior work^17,22,23,36,37^. We applied regularized NMF, which positively weights connectivity patterns that covary, to each individual’s concatenated fMRI time series to identify *k*=17 personalized networks (**Fig 1C**) (**eMethods 5**). A group consensus atlas from past work in an independent dataset^17^ was used as a prior in time series decomposition. This yielded a loading matrix of 17 x 59,412 for each participant, in which 17 corresponds to the number of networks and 59,412 corresponds to the number of cortical vertices. Each value in the loading matrix quantifies the extent to which each vertex belongs to a certain network, a probabilistic network definition that we refer to as PFN topography.

### Statistical Analysis

We leveraged the twin pairs in our sample (254 monozygotic and 334 dizygotic) to calculate heritability of p-factor and PFN vertex-level topography using ACE extended twin design models^43^ controlling for age, sex, and site (as well as mean FD for PFN models) in OpenMX^44^. We computed linear mixed-effects (LME) models to validate univariate associations among p-factor, PRS-F1, and PRS-F2, accounting for fixed effects of age and sex as well as batch effects of family and site (**eMethods 6**).

Next, we trained ridge regression models on the PFN loading matrices of each participant to identify multivariate associations with p-factor, PRS-F1, and PRS-F2 scores. All models included covariates for age, sex, site, and mean FD that were regressed out separately in two half-split subsets based on the ABCD Reproducible Matched Samples^45^. To estimate generalizability across matched subsets, two-fold cross-validation was used: regression models were trained in one subset and tested in an unseen subset, followed by swapping of the training and testing data (**eMethods 7**).

To further interpret multivariate associations between PFN topography and each of our variables of interest, we analyzed the feature weights of our ridge regression models. Feature weights quantify the strength and direction of each network loading’s association with the variable of interest. A positive weight indicates that a network’s loading onto a vertex is associated with a higher score (e.g., high p-factor), and a negative weight indicates that a network’s loading is associated with a lower score (e.g., low p-factor). To account for the covariance structure among features, we applied the Haufe transform^46^ to weights. We averaged Haufe-transformed weights between split-half subsets and interpreted them using three approaches: vertex-level regional importance, network-level importance, and directional network topographies (**eMethods 8**).

## Results

### Overview

Based on 254 monozygotic and 334 dizygotic twin pairs in the ABCD Study® (N=11,873, ages 9-10), we found significant heritability of PFN topography. Transdiagnostic psychopathology indexed by p-factor in youth was associated with polygenic risk for mood-related symptoms and conditions in adults (PRS-F1). Interindividual differences in functional network topography were associated with p-factor, PRS-F1, and PRS-F2 (which captures polygenic risk for psychotic conditions in adults). These multivariate associations were largely driven by the topography of higher-order association networks. We found convergence in individualized topography associated with p-factor and PRS-F1 and divergence in the topography associated with PRS-F2.

### Genetic Effects Underlie P-factor and PFNs

First, we calculated twin-based heritability (*h^2^*) of p-factor and PFN vertex-level topography. A significant proportion of phenotypic variance in p-factor was attributable to additive genetic effects (*h^2^*=0.54, 95% CI [0.41–0.68], p<0.001) (**Fig 2A**). Shared environmental (*C*) and nonshared environmental (*E*) effects accounted for a smaller proportion of p-factor variance (*C*=0.20, 95% CI [0.09–0.30]; *E*=0.26, 95% CI [0.22–0.32]). For PFN topography, we calculated heritability for all non-zero variance PFN loadings and mapped these values thresholded by significance (*p_FDR_*<0.05) to produce 17 network heritability maps (**eFig 2**). We then mapped the maximum heritability of each cortical vertex in terms of its loading on any of the 17 networks (**Fig 2B**). Of these 59,412 heritability estimates, 33,901 (57.1%) were significantly heritable (*p_FDR_*<0.05, mean of maximum *h^2^*=0.35).

**Figure 2.**
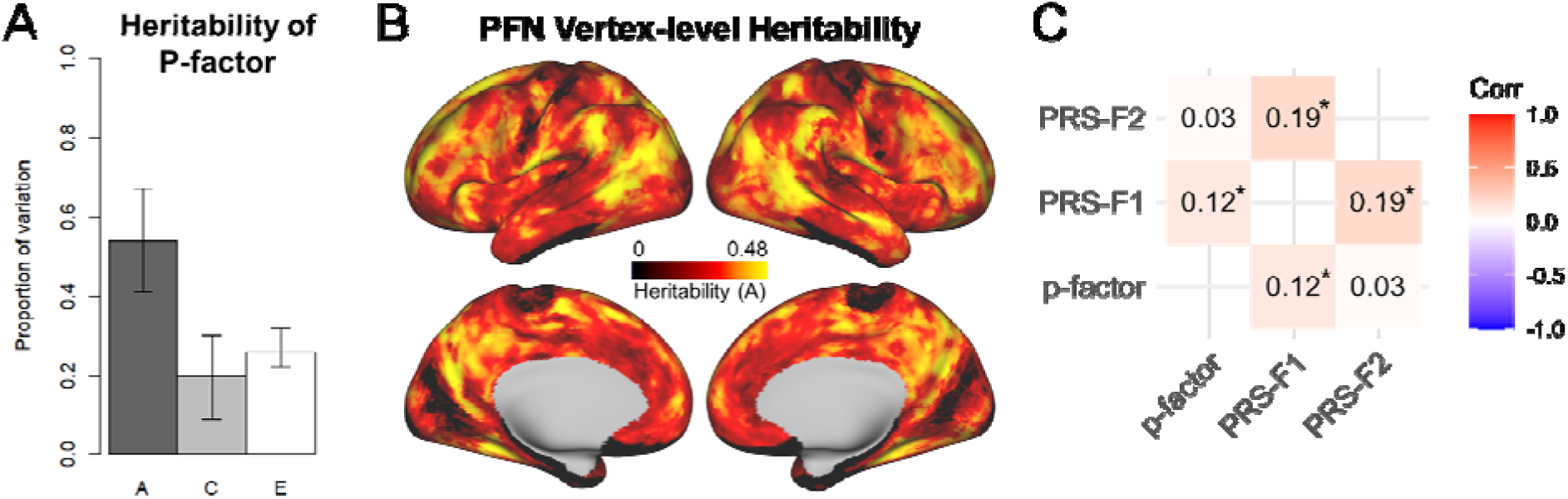
Genetic Effects Underlie Overall Psychopathology and PFN Topography **A.** Leveraging twins included in ABCD, extended twin design models revealed the additive genetic effect (*A*), or heritability (*h^2^*), of p-factor (***h^2^=0.54, 95% CI [0.41–0.68], p<0.001***) (*A* = additive genetic effect, *C* = common environmental effect, *E* = nonshared environmental effect, error bars = 95% CI). **B.** Map showing the maximum heritability of a given cortical vertex across the 17 networks. Out of 59,412 cortical vertices, 33,901 (57.1%) have a maximum heritability that was significantly heritable after false discovery rate (FDR) correction. **C.** Correlation matrix of p-factor, PRS-F1, and PRS-F2 using Pearson correlation coefficients. ***\*p<0.001 based on LME models*.**

Next, we investigated associations between polygenic risk and overall psychopathology by testing correlations among p-factor, PRS-F1, and PRS-F2. We found that p-factor was correlated with PRS-F1 (*r*=0.12, 95% CI [0.09–0.15], *p*<0.001) (**Fig 2C**), validated in an LME model accounting for relevant fixed and batch effects (**eTable 2**; β=0.11, 95% CI [0.09–0.14], *p*<0.001). P-factor was not correlated with PRS-F2 (*r*=0.03, 95% CI [0.00–0.06], *p*=0.05) (**Fig 2C**); our LME model was consistent (**eTable 3**; β=0.02, 95% CI [0.00–0.05], *p*=0.07).

Furthermore, PRS-F2 was not correlated with any of the orthogonal sub-factors (**eFig 3**). Consistent with the correlated factor structure reported^27^ (Fig 1B), PRS-F1 and PRS-F2 were found to be correlated (*r*=0.19, 95% CI [0.17–0.21], *p*<0.001) (**Fig 2C**), validated in our LME model (**eTable 4**; β=0.18, 95% CI [0.15–0.20], *p*<0.001).

### PFN Topography is Associated with Interindividual Differences in P-factor, PRS-F1, PRS-F2

To investigate the multivariate relationship between PFN topography and p-factor, we trained ridge regression models on participant PFN loading matrices and corresponding p-factor scores, followed by testing in unseen data. Model performance, defined as the correlation between the actual p-factor and the model-fit p-factor scores, was significant in both Split-Half-A (*r*=0.12, 95% CI [0.09–0.15]) and Split-Half-B (*r*=0.12, 95% CI [0.09–0.15]) (**Fig 3A**).

**Figure 3.**
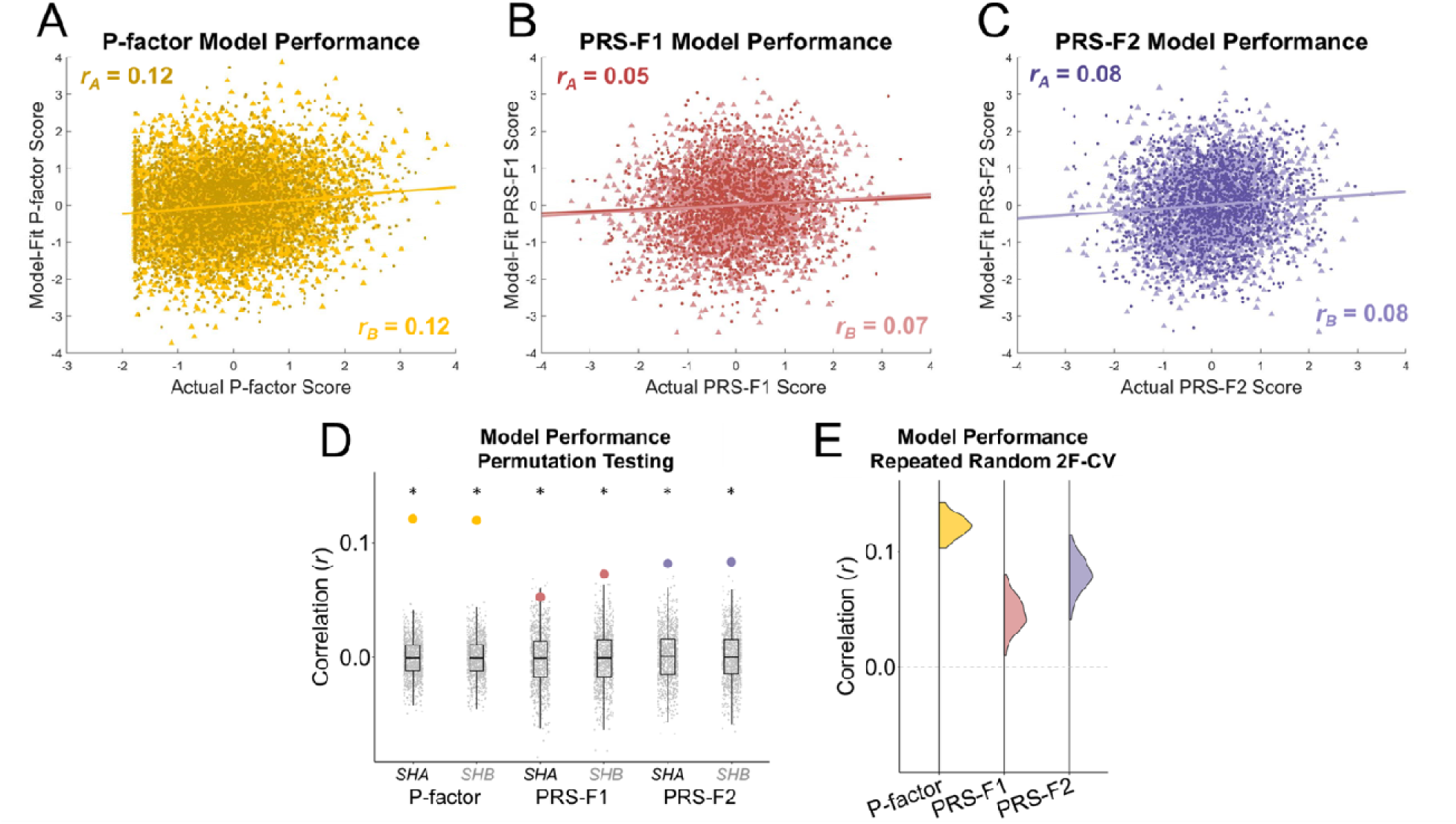
PFN Topography is Associated with Interindividual Differences in P-factor, PRS-F1, and PRS-F2 **A-C.** The multivariate association between PFN topography and p-factor, PRS-F1, and PRS-F2 was assessed using model performance, defined as the correlation between the actual score (p-factor, PRS-F1, or PRS-F2) and the model-fit score produced by PFN-trained ridge regression models. Significant correlations between actual and model-fit scores were found for p-factor (**A**), PRS-F1 (**B**), and PRS-F2 (**C**) in both Split-Half-A (darker scatterplot, ***r_A_***) and Split-Half-B (lighter scatterplot, ***r_B_***) subsets. **D**. Permutation testing (*N*=1000) of model performance revealed that the associations of PFN topography with p-factor, PRS-F1, and PRS-F2 (colored points) were all significantly higher than chance (**\**p<0.01***, box plots show null distributions) (*SHA* = Split-Half-A, *SHB* = Split-Half-B). **E**. Repeated random 2-fold cross-validation (N=100) revealed stability of model performance across randomized sample split-halves.

Then, to investigate the multivariate relationship between PFN topography and psychiatric polygenic risk, we trained and tested ridge regression models on PFN loading matrices and corresponding F1 and F2 polygenic risk scores. Significant correlations between actual and model-fit scores were found for PRS-F1 (Split-Half-A *r*=0.05, 95% CI [0.01–0.10]; Split-Half-B *r*=0.07, 95% CI [0.03–0.12]) (**Fig 3B**) and PRS-F2 (Split-Half-A *r*=0.08, 95% CI [0.04–0.12]; Split-Half-B *r*=0.08, 95% CI [0.04–0.13]) (**Fig 3C**).

Permutation testing revealed that the multivariate associations between PFN topography and each of our variables (p-factor, PRS-F1, and PRS-F2) were all significantly higher than chance (P-factor Split-Half-A *p*<0.001, Split-Half-B *p*<0.001; PRS-F1 Split-Half-A *p*=0.008, Split-Half-B *p*<0.001; PRS-F2 Split-Half-A *p*<0.001, Split-Half-B *p*<0.001) (**Fig 3D**). To ensure robustness to data division, we randomly divided our samples into half-split subsets 100 times, demonstrating stability of PFN model performance on p-factor (mean *r*=0.12), PRS-F1 (mean *r*=0.05), and PRS-F2 (mean *r*=0.08) (**Fig 3E**) across subsets. Scanner manufacturer did not affect model performance (**eFig 4**).

### Cortical Regions Driving PFN Associations Converge Between P-factor and PRS-F1, but Diverge in PRS-F2

To identify important cortical regions driving the association between PFN topography and p-factor, PRS-F1, and PRS-F2, we evaluated the weights of our ridge regression models at the vertex level. Mapping the magnitude of summed weights across networks onto the cortical surface revealed regions driving the association of PFN topography with p-factor (**Fig 4A top**), PRS-F1 (**Fig 4B top**), and PRS-F2 (**Fig 4C top**), a subset of which reached statistical significance (threshold-free cluster enhancement^47^ family-wise error corrected *p*<0.05, **eFig 5**). Unsummed weight maps of individual networks were also derived (**eFigs 6-8**).

**Figure 4.**
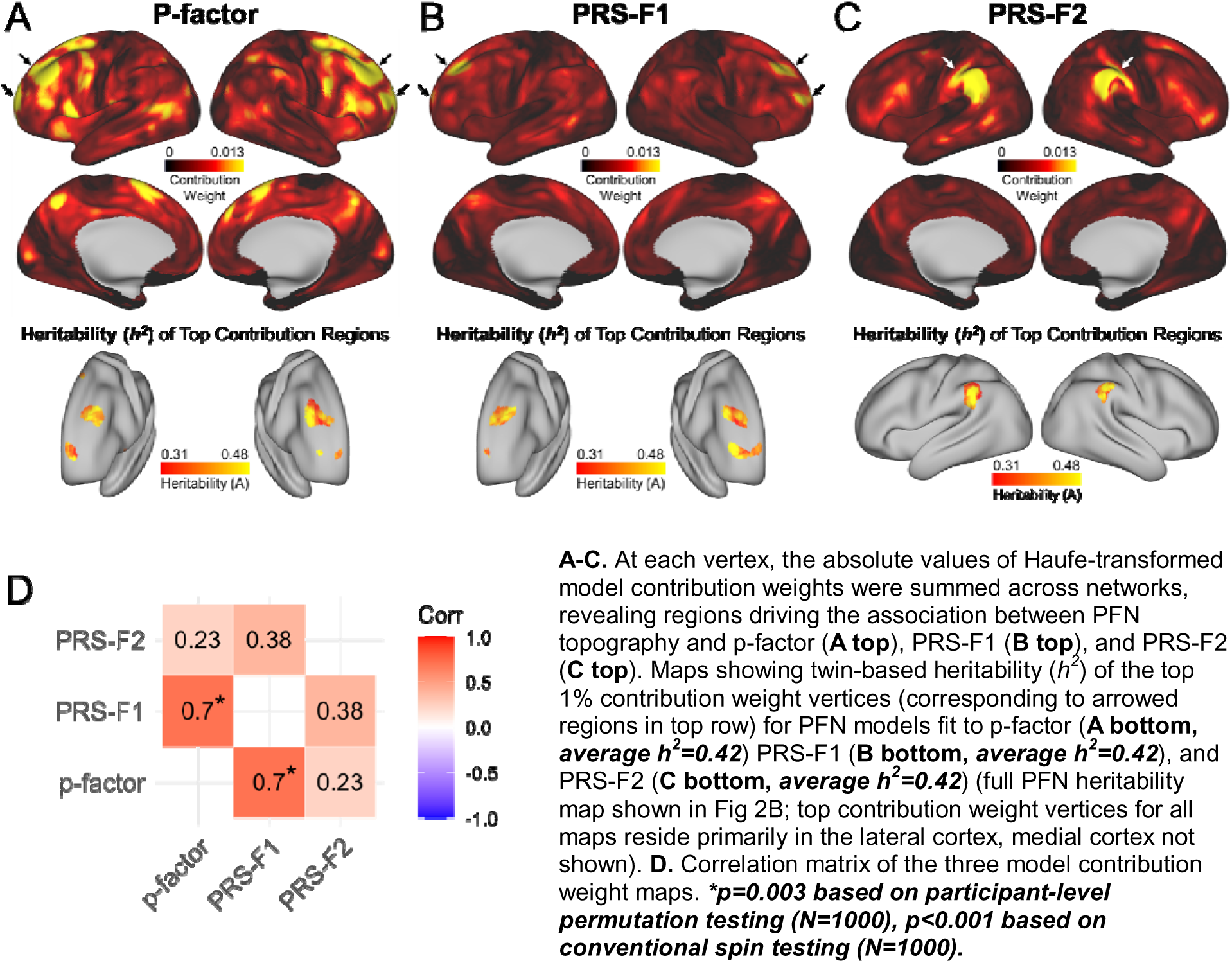
Cortical Regions Driving PFN Associations Converge Between P-factor and PRS-F1, Diverge in PRS-F2 **A-C.** At each vertex, the absolute values of Haufe-transformed model contribution weights were summed across networks, revealing regions driving the association between PFN topography and p-factor (**A top**), PRS-F1 (**B top**), and PRS-F2 (**C top**). Maps showing twin-based heritability (*h^2^*) of the top 1% contribution weight vertices (corresponding to arrowed regions in top row) for PFN models fit to p-factor (**A bottom, *average h^2^=0.42***) PRS-F1 (**B bottom, *average h^2^=0.42***), and PRS-F2 (**C bottom, *average h^2^=0.42***) (full PFN heritability map shown in Fig 2B; top contribution weight vertices for all maps reside primarily in the lateral cortex, medial cortex not shown). **D.** Correlation matrix of the three model contribution weight maps. ***\*p=0.003 based on participant-level permutation testing (N=1000), p<0.001 based on conventional spin testing (N=1000)*.**

Given that p-factor was found to be heritable (Fig 2A) and that PRS-F1 and PRS-F2 are genetic variables, we would expect regions which strongly contribute to PFN associations with these variables to also have significant twin heritability. To test this hypothesis, we investigated the PFN loading heritability (Fig 2B) of vertices with the top 1% contribution weights (i.e., the most important features) for p-factor, PRS-F1, and PRS-F2. The top p-factor and top PRS-F1 associated regions were both found primarily in the dorsolateral and ventromedial prefrontal cortex. Vertices in these regions were nearly all significantly heritable (p-factor cluster 588/594, 99.0%; PRS-F1 cluster 586/594, 98.7%) with substantial average heritability (p-factor *h^2^*=0.42; PRS-F1 *h^2^*=0.42) (**Fig 4A bottom, 4B bottom**). The top PRS-F2 associated vertices, residing primarily in the temporoparietal junction, were nearly all significantly heritable (573/594, 96.5%) with an average heritability of *h^2^*=0.42 (**Fig 4C bottom**).

If the relationship between PRS-F1 and p-factor (Fig 2C) is reflected in functional brain network organization, we might expect shared spatial patterns between the contribution weight maps of PRS-F1 and p-factor. Using participant-level permutation testing, we tested the significance of correlations among all three weight maps. Indeed, we found a strong significant correlation between the weight maps of PRS-F1 and p-factor (*r*=0.70, *p*=0.003) (**Fig 4D**), also significant in conventional spin testing^48^ (*p*<0.001). In contrast, the weight maps of PRS-F2 and p-factor were not found to be significantly correlated (*r*=0.23, permutation testing *p*>0.99, spin testing *p*=0.06) (**Fig 4D**).

### Networks Driving PFN Associations Converge Between P-factor and PRS-F1, but Diverge in PRS-F2

We next sought to identify networks driving the association between PFN topography and p-factor, PRS-F1, and PRS-F2. To do so, we evaluated the aggregate importance of vertices within each network to p-factor (**Fig 5A top**), PRS-F1 (**Fig 5B top**), or PRS-F2 (**Fig 5C top**) models, with each network’s summed weight value normalized by network size. P-factor and PRS-F1 shared the same two most important networks: 15, a frontoparietal (FP) network, and 7, a ventral attention (VA) network. The two most important networks associated with PRS-F2 were divergent: 3, a distinct FP network, and 9, a distinct VA network (**Fig 5D**).

**Figure 5.**
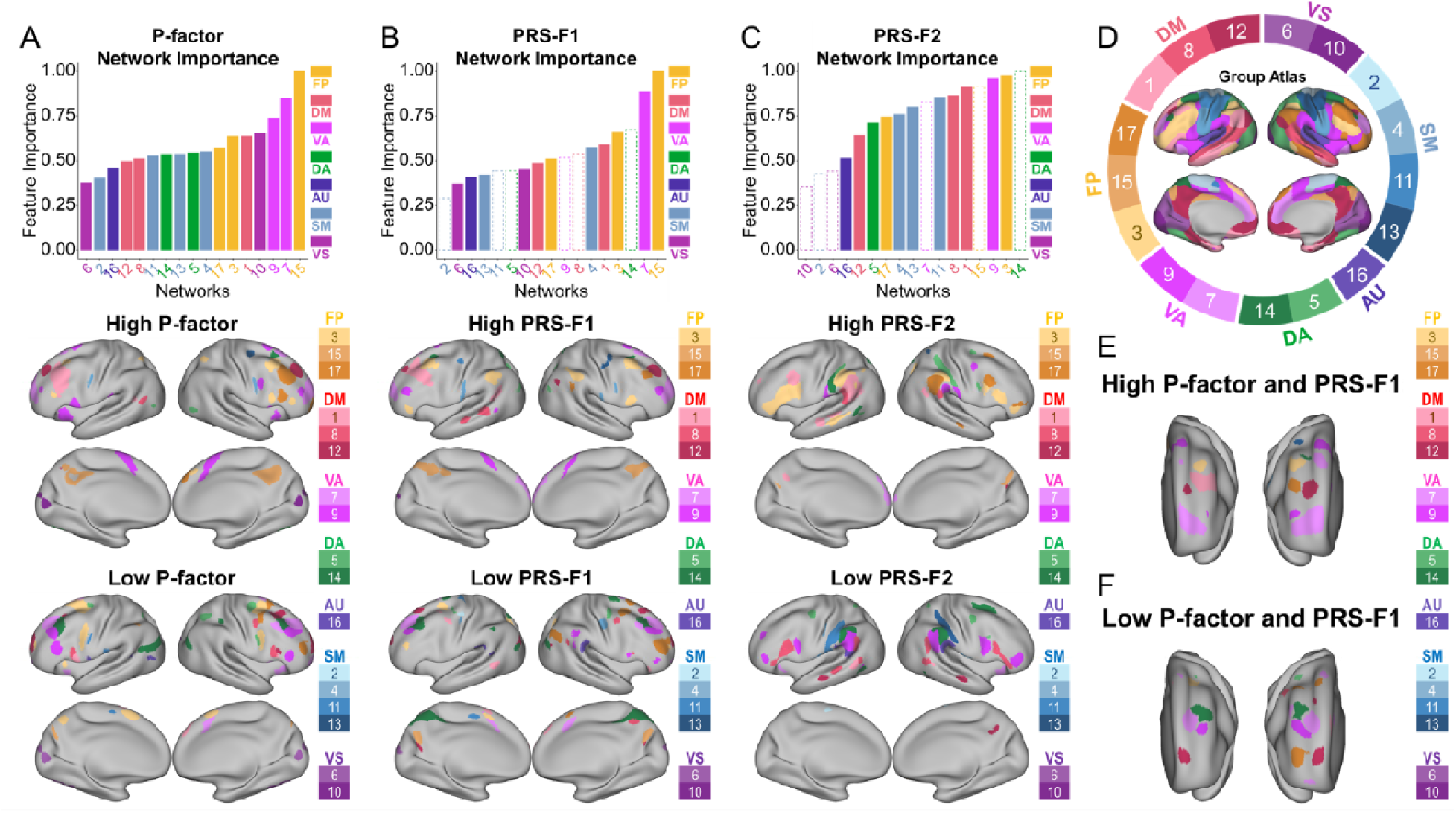
Networks Driving PFN Associations Converge Between P-factor and PRS-F1, Diverge in PRS-F2 **A-C Top.** For each network, the absolute values of Haufe-transformed model contribution weights were summed across vertices, revealing the cumulative feature importance of each network to the multivariate association between PFNs and p-factor (**A top**), PRS-F1 (**B top**), and PRS-F2 (**C top**). Summed weight values were normalized by network size, defined as the number of non-zero vertex loadings within each network of the initial group consensus atlas. The resulting network importance values were normalized to the maximum so that all values are in the range of [0,1] for ease of interpretation. ***Solid bars reflect significance of p<0.05 based on participant-level permutation (N=1000) after FDR correction.*** (FP = frontoparietal, VA = ventral attention, DA = dorsal attention, DM = default mode, AU = auditory, SM = somatomotor, VS = visual). **A-C Middle.** To identify network topography positively associated with our variables of interest, we determined the most positive Haufe-transformed model weight across the 17 networks for each vertex, thresholded to keep the top 10% of most positive weights across the cortical surface, and set a 25mm^2^ cluster threshold. Then, we assigned each of the retained vertices to the network corresponding to each vertex’s most positive weight (network denoted by specific color as shown in legend on the right of each panel). This results in maps of PFN topography associated with high p-factor (**A middle**), high PRS-F1 (**B middle**), and high PRS-F2 (**C middle**). **A-C Bottom.** To identify network topography negatively associated with our variables of interest, we determined the most negative Haufe-transformed model weight across the 17 networks for each vertex, thresholded to keep the top 10% of values (by magnitude) across the cortical surface, and set a 25mm^2^ cluster threshold. Then, we assigned each of the retained vertices to the network corresponding to each vertex’s most negative weight (network denoted by specific color as shown in legend on the right of each panel). This resulted in maps of PFN topography associated with low p-factor (**A bottom**), low PRS-F1 (**B bottom**), and low PRS-F2 (**C bottom**). **D.** Hard parcellation of PFN group atlas for reference. **E**. Conjunction map showing shared topography in prefrontal cortex between high p-factor (A middle) and high PRS-F1 (B middle) maps. **F**. Conjunction map showing shared topography in prefrontal cortex between low p-factor (A middle) and low PRS-F1 (B middle) maps. Whole cortex conjunction maps shown in **eFig 10**.

Next, we sought to understand the specific network topographies underlying these associations. To determine network topography positively associated with p-factor, PRS-F1, and PRS-F2, we assigned vertices corresponding to the top 10% of the most positive weights to their respective networks. This resulted in maps of PFN topography associated with high p-factor (**Fig 5A middle**), high PRS-F1 (**Fig 5B middle**), and high PRS-F2 (**Fig 5C middle**) (top 1% and 5% shown in **eFig 9A-B top**). Notably, there was substantial shared topography between high p-factor and high PRS-F1 maps (2,456/5,941 vertices shared, 41.3%, **eFig 10A top**). Shared positive topography was localized primarily to the prefrontal cortex (**Fig 5E**), including FP networks 3 and 17, default mode (DM) networks 1 and 12, and VA networks 7 and 9. Topography of the high PRS-F2 map diverged from that of p-factor (125/5,941 vertices shared, 2.1%, **eFig 10B top**), with FP networks 3 and 17, DM networks 1 and 8, and VA network 9 localized primarily to the temporoparietal junction (**Fig 5C middle**).

Similarly, we assigned network identity to the top 10% of the most negative weights, resulting in maps of PFN topography associated with low p-factor (**Fig 5A bottom**), low PRS-F1 (**Fig 5B bottom**), and low PRS-F2 (**Fig 5C bottom**) (top 1% and 5% shown in **eFig 9A-B bottom**). There was substantial shared topography between low p-factor and low PRS-F1 maps (1,711/5,941 vertices shared, 28.8%, **eFig 10A bottom**). Shared negative topography was localized primarily to the prefrontal cortex (**Fig 5F**), including FP network 17, DM network 12, VA networks 7 and 9, and dorsal attention (DA) network 14. Topography of the low PRS-F2 map diverged from that of p-factor (269/5,941 vertices shared, 4.5%, **eFig 10B bottom**), with DM network 8, VA network 9, and DA network 14 localized primarily to the temporoparietal junction (**Fig 5C bottom**).

## Discussion

Here, we applied precision functional brain mapping to demonstrate that polygenic risk underlies individual differences in functional network organization during early adolescence. Consistent with our and others’ prior work^23,49^, we showed that PFN topography is associated with psychopathology, and furthermore, that associations of topography with both p-factor and polygenic risk scores are largely driven by association networks. Specific cortical regions and network topographies converged between p-factor and PRS-F1, whereas they diverged in PRS-F2; this aligned with our findings that PRS-F1 was associated with p-factor, whereas PRS-F2 was not. Furthermore, our study replicated past work^34^ showing twin-based heritability of p-factor, and expanded on prior findings of PFN topography heritability during adulthood^26^ by demonstrating its heritability during development. Past work demonstrating excellent test-retest reliability of PFN topography in ABCD (intraclass correlation coefficients: 0.84–0.99)^22^ supports confidence in the robustness of our heritability^50^ and multivariate association results. Together, these findings demonstrate that polygenic risk for transdiagnostic adulthood psychopathology is associated with both p-factor and heritable PFN topography during early adolescence.

Our results showed that PRS-F1, encapsulating liability for common mood disorder symptoms, major depression, and bipolar II disorder in adults, was associated with p-factor during early adolescence. This aligns with prior literature, in which adult-derived polygenic risk scores for depression and neuroticism were associated with general psychopathology in youth^51,52^, including associations described in the ABCD^53,54^. In contrast, PRS-F2, encapsulating liability for schizophrenia, schizoaffective disorder, and bipolar I disorder in adults, was not associated with p-factor or any sub-factor scores in our sample, suggesting that genetic risk for psychotic disorders is not yet phenotypically detectable at this stage of development^55^, at least not in a community sample that is not enriched for clinically high-risk individuals^56^.

Using validated methods of precision functional brain mapping^17,37^ and rigorous cross-validation of our machine learning model, we found that PFN topography is associated with both PRS-F1 and PRS-F2. This association with PRS-F2 implies that genetic risk for psychotic disorders is reflected in the topography of the developing brain before manifesting clinically. The top regions driving the associations of PFN topography with PRS-F1 and PRS-F2 were both highly heritable in twin ACE models, aligning with prior literature on the heritable and polygenic basis of functional brain networks^9,10,57^. However, to our knowledge, this study is the first to demonstrate associations between psychiatric polygenic risk scores and individual-specific functional network topography. It is worth noting that the effect sizes of these associations are small. However, prior work has shown that small samples tend to inflate effect size^58,59^, whereas large samples, as used here, provide more robust estimates.

Cortical regions and network topographies associated with p-factor and PRS-F1 were remarkably similar, primarily residing within the dorsolateral and ventromedial prefrontal cortex. Past literature has found that these regions are altered in neuropsychiatric disorders including mood disorders^60–62^, potentially related to their function in emotional regulation, executive function, and reward-related processes^63,64^. In contrast, cortical regions and network topography associated with PRS-F2 were divergent, residing primarily in the temporoparietal junction. Prior studies have reported that this region is altered in schizophrenia^65–67^, specifically in patients with auditory verbal hallucinations, likely related to dysfunction in language processing^68,69^. Despite this topographical divergence, frontoparietal networks showed maximal importance across p-factor, PRS-F1, and PRS-F2 models, consistent with recent evidence that disruptions in frontoparietal connectivity are common to both affective and psychotic disorders^70^.

Several limitations should be noted. First, PRS-F1 and PRS-F2^27^ do not capture neurodevelopmental disorders such as autism or attention deficit hyperactivity disorder; future precision brain mapping studies should investigate transdiagnostic risk scores that encapsulate a more diverse array of disorders^71^. Second, polygenic risk may comprise only a small proportion of the total heritability of PFN topography^72^; future work should investigate relationships with other genetic factors such as copy number variants^73^. Third, p-factor is a low-dimensional representation of transdiagnostic psychopathology that captures the high comorbidity across disorders^74^ and domain-level measures (e.g. internalizing, externalizing)^75^; however, future studies may show that more nuanced domain-level measures correlate more strongly with genetic risk factors^71,76^. Fourth, this study is cross-sectional, motivating future longitudinal studies leveraging later ABCD imaging time points to gain further causal insight.

Here, we sought to uncover biological mechanisms underlying the emergence of mental illness using precision functional mapping. Together, the present results are a meaningful step towards understanding the genetic and functional drivers of transdiagnostic psychopathology in early adolescence.

## Data Availability

Data used in the preparation of this article were obtained from the Adolescent Brain Cognitive Development Study^®^ (https://abcdstudy.org), held in the NIMH Data Archive (NDA). Only researchers with an approved NDA Data Use Certification (DUC) may obtain ABCD Study data.

## Acknowledgments

AAB had full access to all the data in the study and takes responsibility for the integrity of the data and the accuracy of the data analysis.

KYS was supported by 1F30MH138048-01, KYS, EK, ZS, JS, and AAB were supported by 5R01MH132934-02, KYS, ASK, and TDS were supported by 5R01MH113550-07 and 5R01MH120482-03, LA and LMS were supported by 5U01MH119690, TTM was supported by 1K08MH135343, HL, YF, and TDS were supported by 5R01EB022573, and ASK was supported by 5T32MH019112-32 and 1L30MH131061-01 from the National Institutes of Health. The funder had no role in the design and conduct of the study; collection, management, analysis, and interpretation of the data; preparation, review, or approval of the manuscript; and decision to submit the manuscript for publication.

AAB has received consulting income from Octave Bioscience. JS and AAB hold shares in and JS is a director of Centile Bioscience Inc.

