## Supplemental Materials for "Polygenic Risk, Psychopathology, and Personalized Functional Brain Network Topography in Adolescence"

**eMethods 1. Participants**

**eMethods 2. P-factor Derivation**

**eMethods 3. F1 and F2 Polygenic Risk Score Derivation**

**eMethods 4. Image Processing**

**eMethods 5. Regularized Non-negative Matrix Factorization (NMF)**

**eMethods 6. Linear Mixed-Effects (LME) Models**

**eMethods 7. Ridge Regression Models and Validation**

**eMethods 8. Ridge Regression Weight Analysis**

**eTable 1. Confirmatory Bifactor Model of ABCD Clinical Items**

**eFig 1. Correlation between P-factor Scores from Longitudinal vs. Cross-sectional Bifactor Model**

**eTable 2. P-factor and PRS-F1 LME model**

**eTable 3. P-factor and PRS-F2 LME model**

**eTable 4. PRS-F1 and PRS-F2 LME model**

**eFig 2. Significant Heritabilities of PFN vertex loadings across 17 networks**

**eFig 3. PRS-F1 and PRS-F2 Correlations with the 8 Sub-factor Scores**

**eFig 4. Ridge Regression Models with inclusion of Scanner Manufacturer as a Covariate**

**eFig 5. Significance Testing of Cortical Regions Driving PFN Associations with P-factor, PRS-F1, and PRS-F2**

**eFig 6. P-factor Weight Maps**

**eFig 7. PRS-F1 Weight Maps**

**eFig 8. PRS-F2 Weight Maps**

**eFig 9. Top 5% and 1% Directional Network Topography Maps**

**eFig 10. Conjunction Maps of Top 10% Directional Network Topographies**

This supplementary material has been provided by the authors to give readers additional information about their work.

**eMethods 1. Participants**

Exclusion criteria of the Adolescent Brain Cognitive Developmental (ABCD) Study℠ included severe sensory, intellectual, medical, or neurological issues that would interfere with the youth’s ability to comply with the study protocol, as well as MRI scanner contraindications^1^. In our imaging analyses, we additionally excluded participants with incomplete data or excessive head motion, yielding a sample of *N* = 7,459 youths. Furthermore, in our imaging analyses including polygenic risk scores of F1 and F2, we used a European ancestry subsample of *N* = 3,982 youths, as the original Mallard et al. 2022 multivariate genome-wide association study (GWAS)^2^ was performed in a European ancestry sample^3^.

**eMethods 2. P-factor Derivation**

At the baseline and year 2 time points of the ABCD Study**^®^**, youths and caregivers completed a consistent series of questionnaires assessing the youth’s mental health, which include the Kiddie Schedule for Affective Disorders and Schizophrenia (KSADS-5), Prodromal Questionnaire–Brief Child Version (PQ-BC), Child Behavior Checklist (CBCL), and General Behavior Inventory 10 item Mania Scale (GBMI)^4^.

To capture a participant’s overall psychopathology, we used a longitudinal bifactor model, in which each mental health interview item loads onto both a general factor (p-factor) and one of multiple orthogonal sub-factors. Although our analyses only used the baseline factor scores, this longitudinal model was used to optimize signal-to-noise ratio via inclusion of twofold more data compared to a cross-sectional bifactor. Analysis followed a split-sample exploratory-to-confirmatory procedure similar to that used in Moore et al. 2020^5^. Data were first split randomly into exploratory and confirmatory subsamples by family, such that siblings were guaranteed to be in the same sub-sample to avoid leakage. In the exploratory sub-sample (*N*=5,938; independent measures = 11,119), item-factor analysis of 178 items was performed by way of exploratory structural equation modeling (ESEM)^6^ using the mean- and variance-adjusted weighted least-squares (wlsmv) estimator^7,8^ and oblimin rotation, extracting eight factors, in Mplus v8.4^9^. The analysis included stratification by site (22 sites, one is no longer active), clustering by family (5,421 families), and grouping by time point (two time points, baseline, ages 9-10, and year 2, ages 11-12) with the constraint that model loadings and thresholds had to be invariant across time. The number of factors to extract was determined by a combination of interpretability and visual examination of the scree plot. Empirical methods for determining the number of factors, including the minimum average partial method^10^ and parallel analysis with Glorfeld correction^11^, suggested clear over-extractions (19 and 39 factors, respectively). Model fit was assessed using the acceptability cutoffs suggested by Hu and Bentler (1999)^12^ via the root mean-square error of approximation (RMSEA; acceptable = 0.06), comparative fit index (CFI; acceptable = 0.95), Tucker-Lewis index (TLI; acceptable = 0.95), and standardized root mean-square residual (SRMR; acceptable = 0.08).

Next, in the confirmatory sub-sample (*N* = 5,938; independent measures = 11,168), confirmatory factor analysis (CFA) was performed using the item configuration suggested by the exploratory analysis, omitting items that loaded < 0.40 on their respective factors, leaving 125 items for the CFA. In addition to the eight factors suggested by the ESEM, a general factor of psychopathology (p-factor) was also included, making a bifactor configuration^13,14^ where all sub-factors are orthogonal to the general factor and to each other. Consistent with the ESEM, the CFA included stratification by site (22 sites), clustering by family (5,431 families), and grouping by time point (two time points) with the constraint that model loadings and thresholds had to be invariant across time. Model fit was assessed using the indices and cutoff criteria listed above.

Fit of the exploratory (ESEM) model was acceptable (CFI = 0.97; RMSEA = 0.011; SRMR = 0.046), with 125 out of the 178 items having absolute loadings > 0.40. These 125 items were then carried forward to the bifactor CFA in the testing sub-sample. Fit of the confirmatory model was acceptable (CFI = 0.95; RMSEA = 0.017; SRMR = 0.063) and with that acceptable fit confirmed, the CFA was re-estimated on the full sample (training and testing combined) to obtain factor scores. The final bifactor model consisted of loadings onto the p-factor as well as 8 orthogonal sub-factors, which correspond to items broadly associated with psychosis (PSY), mania (MAN), oppositional defiance (OPP), internalizing symptoms (INT), attention-deficiency and hyperactivity (ADH), somatic symptoms (SOM), disordered eating (DSE), and post-traumatic stress (PTS) (**eTable 1**). Bifactor-specific reliability and generalizability indices were all above normally acceptable cutoffs^15^ for the general factor: omegaH = 0.83, H = 0.98, factor determinacy = 0.97, percent uncontaminated correlations = 0.82. We used the baseline p-factor score for our analyses. As a sensitivity analysis, we derived a cross-sectional bifactor model using baseline time point data with the same factor structure as the longitudinal model, albeit with different item loading values. We found the resulting p-factor scores from the cross-sectional bifactor to be highly correlated to baseline p-factor from the longitudinal bifactor (r=0.9997, **eFig 1**).

**eMethods 3.** **F1 and F2 Polygenic Risk Score Derivation**

At the baseline time point of the ABCD study^16^, saliva samples were collected, and DNA was genotyped on the Affymetrix Smokescreen array. Before imputation, PLINK 1.9^17^ was used to remove SNPs with >5% missingness, samples with more than 10% missingness, and samples in which genotyped sex did not match reported sex. Genotypes were phased (Eagle v.2.4) and imputed to the 1000 Genomes Project Phase 3v5 reference panel^18^ using Minimac 4 via the Michigan Imputation Server^19^. After imputation, only sites with imputation quality *R^2^* ≥ 0.7 and MAF ≥ 0.01 were retained.

Latent genetic factors F1 and F2 were derived from a multivariate GWAS^2^ based on European ancestry adults (UK Biobank^20^, Psychiatric Genomics Consortium^21–23^, and other cohorts^21,22^). Mallard et al. 2022^2^ was chosen over other multivariate GWAS approaches^24^ as its corresponding summary statistics were publicly available. Another advantage of the Mallard et al. 2022 approach is its relatively balanced statistical design, which decreases potential bias due to disparities between datasets^2^. To account for population-level variability in allele frequencies and linkage disequilibrium, ancestry must be considered^25^. Multidimensional scaling (MDS) of the imputed ABCD genotypes was conducted using KING v.2.2.4^26^ to identify the top 10 ancestry principal components (PCs). These PCs were then projected onto the 1000 Genomes Project Phase 3v5 PC space and a support vector machine algorithm^27^ was used to infer genetic ancestry and identify a European (EUR) ancestry subsample of ABCD. A second round of unprojected MDS was then performed within the EUR-ancestry subsample to produce the top 10 PCs for use in ancestry-specific analyses.

In the EUR-ancestry subsample, polygenic risk scores of F1 and F2 were calculated using PRC-CS and PLINK. PRS-CS is a Bayesian polygenic prediction method that applies a continuous shrinkage prior to SNP effect sizes^28^. Polygenic scores were calculated using PLINK by summing all included variants weighted by the inferred posterior effect size for that allele and converting the value to a Z-score for each participant within the prediction dataset. Standardized PRS were then adjusted by regressing out the first ten EUR-ancestry PCs.

**eMethods 4. Image Processing**

As described in previous work^29,30^, imaging data was preprocessed according to the ABCD-BIDS pipeline^31^**.** Briefly, the pipeline includes preprocessing of structural T1 MRI data with Advanced Normalization Tools (ANTs), FreeSurfer segmentation and surface registration, as well as volume registration using FSL FLIRT rigid-body transformation. Functional MRI preprocessing included (1) de-meaning and de-trending all fMRI data with respect to time; (2) denoising using a general linear model including regressors for signal and movement; (3) bandpass filtering between 0.008 and 0.09 Hz; (4) applying a respiratory motion filter, and (5) applying motion censoring (frames exceeding a framewise displacement (FD) threshold of 0.2mm or failing to pass outlier detection at +/− 3 standard deviations were discarded).

Consistent with previous work^29,30^, we concatenated the time series data from up to four resting-state scans and three task-based scans (Monetary Incentive Delay Task, Stop-Signal Task, and Emotional N-Back Task) to maximize the available data for our analyses, yielding a maximum concatenated length of 29 minutes and 36 seconds. Participants with fewer than 600 remaining TRs after motion censoring or who failed to pass ABCD quality control for their T1 or resting-state fMRI scan were excluded. We calculated mean framewise displacement (mean FD) for each participant’s concatenated time series across runs to include as a motion covariate in our analyses.

**eMethods 5. Regularized Non-negative Matrix Factorization (NMF)**

As previously ^29,30,32,33^, we used a regularized NMF method^34^ to derive individualized functional networks. As NMF requires nonnegative input, we re-scaled data by shifting time courses of each vertex linearly to ensure positive values. To avoid features in greater numeric ranges dominating those in smaller numeric ranges, the time course was further normalized by its maximum value so that all values are in the range of [0,1]. For this study, identical parameter settings were used as in prior PFN studies.

Spatial correspondence across subjects was ensured by using a group consensus atlas from a previously published study in an independent dataset as an initialization for network definition^32^. Briefly, to derive this group consensus atlas, group-level decomposition was performed 50 times on independent subsets of 100 randomly selected subjects. Inter-network similarity among the 50 decomposition results was calculated and normalized-cuts based spectral clustering was applied to group networks into 17 clusters. For each cluster, the network with the highest overall similarity to the other networks within the same cluster was selected as the representative network. The resulting group-level network loading matrix was transformed from *fsaverage5* space to *fslr* space using Connectome Workbench^35^, with the resultant matrix having 17 columns and 59,412 rows. To derive individualized functional networks, we applied NMF to individual fMRI time series, using the previously-derived group consensus atlas as a prior to ensure inter-individual correspondence as well as a data locality regularization term to improve robustness to imaging noise.

**eMethods 6. Linear Mixed-Effects (LME) Models**

We computed linear mixed-effects (LME) models using the package ‘lme4’^36^ in R version 4.4.0 to assess univariate associations between p-factor and polygenic risk scores PRS-F1 and PRS-F2, while accounting for fixed and random parameters (**eTables 2-4**). Parameters included fixed effects for age and biological sex, as well as a random intercept for family to account for siblings included within our sample. Data were harmonized across sites via neuroCombat^37,38^ prior to linear mixed-effects analysis.

**eMethods 7. Ridge Regression Models and Validation**

To assess the multivariate associations between PFN topography with overall psychopathology and polygenic risk, we trained ridge regression models on PFN vertex-wise network loading matrices of each participant fit to p-factor, PRS-F1, and PRS-F2 using ‘RidgeCV’ function within the ‘scikit-learn’ library in Python, the same approach as used in prior work^29,30^. All three models included covariates for age, biological sex, site, and head motion (mean FD) that were regressed out separately in split-half subsets based on the ABCD Reproducible Matched Samples^39^ (p-factor model *N* = 3,711 and *N* = 3,748; PRS models *N* = 1,978 and *N* = 2,004). As a sensitivity analysis, we included ridge regression models with an additional covariate of scanner manufacturer (**eFig 4**).

Regression models were then trained and tested using nested cross-validation (CV), with the inner CV determining the optimal tuning parameter (λ) for ridge regression and the outer CV estimating the generalizability of the model. For the inner CV, λ values across a range (1, 10, 100, 500, 1000, 5000, 10000, 15000, 20000) were evaluated in each subset using efficient leave-one-out cross-validation, in which the negative mean squared error is calculated to score and determine the optimal λ.

For the outer CV, model performance of the ridge regression was assessed, training on one subset using its respective optimal λ and then testing on the other, as well as vice versa. Model performance is defined in both split-half subsets as the correlation *r* between the observed and model-fit score. Although these are predictive models, it is important to note that we are not using model performance results to make claims about causal directionality, instead using ridge regression as a tool to understand the multivariate associations between the high dimensional PFN topography and our variables of interest: p-factor, PRS-F1, and PRS-F2.

To test the significance of these multivariate associations, we used participant-level permutation of scores (*N* = 1000 permutations), randomly shuffling each participant’s p-factor, PRS-F1, and PRS-F2 scores, then training and testing ridge regression as described above to generate a null distribution of model performance for each model in each half-split subset. Furthermore, to ensure that our matched split-half subsets did not bias our results, we performed repeated random cross-validation (*N* = 100 iterations), each time randomly splitting the sample to generate a distribution of model performance for each score.

**eMethods 8. Ridge Regression Weight Analysis**

To improve interpretability and to account for the covariance structure among features^40^, we first applied the Haufe transform to weights, calculating the covariance between the model-fit score and each PFN loading value across participants and then normalizing by the variance of the actual score. Then, to summarize shared important features across split-half subsets, we averaged the Haufe-transformed weights between the two subsets. We interpreted these averaged Haufe-transformed weights using three different approaches: vertex-level importance, network-level importance, and directional network topographies.

For the vertex-level interpretation, we summed the absolute value of Haufe-transformed weights across networks for each vertex, resulting in 59,412 values, each corresponding to the absolute association of a particular vertex with p-factor, PRS-F1, or PRS-F2. We chose to take the absolute value of weights for this analysis to capture the total contribution of a vertex’s network loadings to model fit, regardless of the directionality of association with specific networks (positive or negative). Statistical significance of the cortical regions was tested using threshold-free cluster enhancement (TFCE) of the summed absolute Haufe weights and a participant-level permutation approach (*N* = 1000 permutations)^41^, with the resulting family-wise-error-corrected p-values used to generate thresholded weight maps (**eFig 5**). Unsummed, directional weight maps of each network for each model are shown in **eFig 6-8**. To determine the shared spatial patterning of vertices associated with p-factor, PRS-F1, and PRS-F2, we tested the correlations between the maps using neuromaps^42^, testing significance using a participant-level permutation approach (*N* = 1000 permutations), randomly shuffling each participant’s scores, then training and testing ridge regression models as described above to generate null distributions of map correlations. We also used spatial randomization spin testing^43^ (*N* = 1000 randomizations) to test significance of weight map correlations.

For the network-level interpretation, we summed the absolute value of Haufe-transformed weights across vertices within each network, resulting in 17 summed weights values, each corresponding to the absolute association of a particular network with p-factor, PRS-F1, or PRS-F2. To determine network-level importance irrespective of the cortical surface area of a given network, summed weights values were normalized by network size, defined as the number of non-zero vertex loadings within each network of the initial PFN group consensus atlas. The resulting network importance values were normalized to the maximum so that all values are in the range of [0,1] for ease of interpretation. To test the significance of network importance, we used participant-level permutation of scores (*N* = 1000 permutations), randomly shuffling each participant’s p-factor, PRS-F1, and PRS-F2 scores, then training and testing ridge regression models as described above to generate null distributions of absolute summed network weight values.

To interpret directional network topography, we identified the most positive and the most negative Haufe-transformed model weight across the 17 networks for each vertex. Then, we thresholded to keep the top 10% most positive and the top 10% most negative weights, and subsequently set a 25mm^2^ cluster threshold on each map. Finally, for each map, we assigned each of the retained vertices to the network corresponding to the vertex’s identified most positive or negative weight, allowing us to understand the networks driving cortical region associations and their directionality. This results in a total of 6 maps: PFN topography associated with high and low p-factor, high and low PRS-F1, and high and low PRS-F2. Additional directional network topographies maps were derived for top 1% and 5% most positive and negative weights (**eFig 9**). Conjunction maps between top 10% directional network topographies were derived, showing the degree to which networks overlap between directional maps of p-factor and those of PRS-F1 and PRS-F2 (**eFig 10**).

| **eTable 1. Confirmatory Bifactor Model of ABCD Clinical Items** | | | | | | | | | | | |
| --- | --- | --- | --- | --- | --- | --- | --- | --- | --- | --- | --- |
| **Item** | **ABCD Instrument** | **Item Description** | **P-factor** | **PSY** | **MAN** | **OPP** | **INT** | **ADH** | **SOM** | **DSE** | **PTS** |
| prodromal_14_y | ABCD Youth Prodromal Questionnaire–Brief Child Version (PQ-BC)  ABCD Youth Prodromal Questionnaire–Brief Child Version (PQ-BC) | Did you feel confused because something you experienced didn't seem real or it seemed imaginary to you? | 0.154 | 0.680 |  |  |  |  |  |  |  |
| prodromal_20_y |  | Have you seen things that other people can't see or don't seem to see? | 0.197 | 0.718 |  |  |  |  |  |  |  |
| prodromal_10_y |  | Did you lose concentration because you noticed sounds in the distance that you usually don't hear? | 0.168 | 0.708 |  |  |  |  |  |  |  |
| prodromal_19_y |  | Did you suddenly start to see unusual things that you never saw before like flashes, flames, blinding light, or shapes floating in front of you? | 0.173 | 0.714 |  |  |  |  |  |  |  |
| prodromal_21_y |  | Did you suddenly start to notice that people sometimes had a hard time understanding what you were saying, even though they used to understand you well? | 0.144 | 0.673 |  |  |  |  |  |  |  |
| prodromal_11_y |  | Although you could not see anything or anyone, did you suddenly start to feel that an invisible energy, creature, or some person was around you? | 0.167 | 0.660 |  |  |  |  |  |  |  |
| prodromal_5_y |  | Did you feel that someone else, who is not you, has taken control over the private, personal, thoughts or ideas inside your head? | 0.188 | 0.647 |  |  |  |  |  |  |  |
| prodromal_12_y |  | Did you start to worry at times that your mind was trying to trick you or was not working right? | 0.181 | 0.624 |  |  |  |  |  |  |  |
| prodromal_16_y |  | Did you feel that parts of your body had suddenly changed or worked differently than before; like your legs had suddenly turned to something else or your nose could suddenly smell things you'd never actually smelled before? | 0.146 | 0.674 |  |  |  |  |  |  |  |
| prodromal_18_y |  | Did you feel that other people might want something bad to happen to you or that you could not trust other people? | 0.187 | 0.654 |  |  |  |  |  |  |  |
| prodromal_4_y |  | Did you feel like you had special, unusual powers like you could make things happen by magic, or that you could magically know what was inside another person's mind, or magically know what was going to happen in the future when other people could not? | 0.150 | 0.653 |  |  |  |  |  |  |  |
| prodromal_7_y |  | Did you ever feel very certain that you have very special abilities or magical talents that other people do not have? | 0.138 | 0.652 |  |  |  |  |  |  |  |
| prodromal_13_y |  | Did you feel that the world is not real, you are not real, or that you are dead? | 0.175 | 0.633 |  |  |  |  |  |  |  |
| prodromal_17_y |  | Did you feel that sometimes your thoughts were so strong you could almost hear them, as if another person, NOT you, spoke them? | 0.165 | 0.645 |  |  |  |  |  |  |  |
| prodromal_8_y |  | Did you suddenly feel that you could not trust other people because they seemed to be watching you or talking about you in an unfriendly way? | 0.152 | 0.646 |  |  |  |  |  |  |  |
| prodromal_2_y |  | Did you hear strange sounds that you never noticed before like banging, clicking, hissing, clapping, or ringing in your ears? | 0.128 | 0.639 |  |  |  |  |  |  |  |
| prodromal_6_y |  | Did you suddenly find it hard to figure out how to say something quickly and easily so that other people would understand what you meant? | 0.098 | 0.618 |  |  |  |  |  |  |  |
| prodromal_1_y |  | Did places that you know well, such as your bedroom, or other rooms in your home, your classroom or school yard, suddenly seem weird, strange or confusing to you; like not the real world? | 0.206 | 0.612 |  |  |  |  |  |  |  |
| prodromal_3_y |  | Do things that you see appear different from the way they usually do (brighter or duller, larger or smaller, or changed in some other way)? | 0.127 | 0.618 |  |  |  |  |  |  |  |
| prodromal_9_y |  | Do you sometimes get strange feelings on or just beneath your skin, like bugs crawling? | 0.160 | 0.610 |  |  |  |  |  |  |  |
| prodromal_15_y |  | Did you honestly believe in things that other people would say are unusual or weird? | 0.126 | 0.527 |  |  |  |  |  |  |  |
| Anhedonia_y | ABCD Youth Diagnostic Interview for DSM-5 5 (KSADS-5) | Symptom - Anhedonia (Past or Present) | 0.189 | 0.541 |  |  |  |  |  |  |  |
| Irritability_y |  | Symptom - Irritability (Past or Present) | 0.222 | 0.529 |  |  |  |  |  |  |  |
| DepressedMood_y |  | Symptom - Depressed Mood (Past or Present) | 0.219 | 0.494 |  |  |  |  |  |  |  |
| ExcessiveWorriesMoreDaysThanNot_y |  | Symptom - Excessive worries more days than not (Past or Present) | 0.194 | 0.495 |  |  |  |  |  |  |  |
| SuicidalIdeation_y |  | Symptom - Suicidal Ideation (Past or Present) | 0.316 | 0.516 |  |  |  |  |  |  |  |
| SelfInjuriousBehavior_y |  | Symptom - Self injurious behavior  (Past or Present) | 0.279 | 0.453 |  |  |  |  |  |  |  |
| Insomnia_y |  | Symptom - Insomnia (Past or Present) | 0.185 | 0.497 |  |  |  |  |  |  |  |
| WishesBetterOffDead_y |  | Symptom - Wishes/Better off dead  (Past or Present) | 0.287 | 0.504 |  |  |  |  |  |  |  |
| FearOfSocialSituations_y |  | Symptom - Fear of Social Situations  (Past or Present) | 0.181 | 0.507 |  |  |  |  |  |  |  |
| DecreasedNeedForSleep_y |  | Symptom - Decreased Need for Sleep  (Past or Present) | 0.161 | 0.477 |  |  |  |  |  |  |  |
| ElevatedMood_y |  | Symptom - Elevated Mood (Past or Present) | 0.093 | 0.426 |  |  |  |  |  |  |  |
| gen_child_behav_4 | ABCD Parent General Behavior Inventory 10 item Mania Scale (GBMI)  ABCD Parent General Behavior Inventory 10 item Mania Scale (GBMI) | Has your child had periods of extreme happiness and intense energy that last several days or more when he/she also felt more anxious or tense (jittery, nervous, uptight) than usual (other than relates to the menstrual cycle)? | 0.636 |  | 0.652 |  |  |  |  |  |  |
| gen_child_behav_2 |  | Have there been periods of several days or more when your child's friends or other family members told you that your child seemed unusually happy or high - clearly different from his/her usual self or from a typical good mood? | 0.546 |  | 0.625 |  |  |  |  |  |  |
| gen_child_behav_5 |  | Have there been times of several days or more when, although your child was feeling unusually happy and intensely energetic (clearly more than his/her usual self), he/she also had to struggle very hard to control inner feelings of rage or an urge to smash or destroy things? | 0.713 |  | 0.514 |  |  |  |  |  |  |
| gen_child_behav_1 |  | Has your child experienced periods of several days or more when, although he/she was feeling unusually happy and intensely energetic (clearly more than your child's usual self), he/she was also physically restless, unable to sit still, and had to keep moving or jumping from one activity to another? | 0.651 |  | 0.538 |  |  |  |  |  |  |
| gen_child_behav_6 |  | Has your child had periods of extreme happiness and intense energy (clearly more than his/her normal self) when, for several days or more, it took him/her over an hour to get to sleep at night? | 0.590 |  | 0.598 |  |  |  |  |  |  |
| gen_child_behav_9 |  | Have there been periods when, although your child was feeling unusually happy and intensely energetic, almost everything got on his/her nerves and made him/her irritable or angry (other than related to the menstrual cycle)? | 0.703 |  | 0.503 |  |  |  |  |  |  |
| gen_child_behav_8 |  | Has your child had periods lasting several days or more when he/she felt depressed or irritable, and then other periods of several days or more when he/she felt extremely high, elated, and overflowing with energy? | 0.730 |  | 0.490 |  |  |  |  |  |  |
| gen_child_behav_7 |  | Have you ever found that your child's feelings or energy are generally up or down, but rarely in the middle? | 0.730 |  | 0.426 |  |  |  |  |  |  |
| gen_child_behav_3 |  | Has your child's mood or energy shifted rapidly back and forth from happy to sad or high to low? | 0.700 |  | 0.401 |  |  |  |  |  |  |
| gen_child_behav_10 |  | Has your child had times when his/her thoughts and ideas came so fast that he/she couldn't get them all out, or they came so quickly others complained that they couldn't keep up with your child's ideas? | 0.623 |  | 0.368 |  |  |  |  |  |  |
| ElevatedMood | ABCD Parent Diagnostic Interview for DSM-5 Full (KSADS-5) | Symptom - Elevated Mood (Past or Present) | 0.210 |  | 0.489 |  |  |  |  |  |  |
| DecreasedNeedForSleep |  | Symptom - Decreased Need for Sleep  (Past or Present) | 0.453 |  | 0.308 |  |  |  |  |  |  |
| cbcl_q22_p | ABCD Parent Child Behavior Checklist (CBCL)  ABCD Parent Child Behavior Checklist (CBCL) | Disobedient at home | 0.646 |  |  | 0.547 |  |  |  |  |  |
| cbcl_q57_p |  | Physically attacks people | 0.586 |  |  | 0.508 |  |  |  |  |  |
| cbcl_q03_p |  | Argues a lot | 0.640 |  |  | 0.480 |  |  |  |  |  |
| cbcl_q97_p |  | Threatens people | 0.671 |  |  | 0.498 |  |  |  |  |  |
| cbcl_q16_p |  | Cruelty, bullying, or meanness to others | 0.585 |  |  | 0.511 |  |  |  |  |  |
| cbcl_q28_p |  | Breaks rules at home, school or elsewhere | 0.680 |  |  | 0.503 |  |  |  |  |  |
| cbcl_q95_p |  | Temper tantrums or hot temper | 0.662 |  |  | 0.387 |  |  |  |  |  |
| cbcl_q37_p |  | Gets in many fights | 0.615 |  |  | 0.436 |  |  |  |  |  |
| cbcl_q21_p |  | Destroys things belonging to their family or others | 0.688 |  |  | 0.446 |  |  |  |  |  |
| cbcl_q86_p |  | Stubborn, sullen, or irritable | 0.683 |  |  | 0.269 |  |  |  |  |  |
| cbcl_q26_p |  | Doesn't seem to feel guilty after misbehaving | 0.623 |  |  | 0.409 |  |  |  |  |  |
| cbcl_q43_p |  | Lying or cheating | 0.597 |  |  | 0.412 |  |  |  |  |  |
| cbcl_q68_p |  | Screams a lot | 0.651 |  |  | 0.333 |  |  |  |  |  |
| cbcl_q23_p |  | Disobedient at school | 0.649 |  |  | 0.406 |  |  |  |  |  |
| cbcl_q94_p |  | Teases a lot | 0.563 |  |  | 0.388 |  |  |  |  |  |
| cbcl_q90_p |  | Swearing or obscene language | 0.564 |  |  | 0.354 |  |  |  |  |  |
| cbcl_q81_p |  | Steals at home | 0.594 |  |  | 0.417 |  |  |  |  |  |
| cbcl_q20_p |  | Destroys their own things | 0.712 |  |  | 0.343 |  |  |  |  |  |
| cbcl_q27_p |  | Easily jealous | 0.628 |  |  | 0.198 |  |  |  |  |  |
| cbcl_q25_p |  | Doesn't get along with other kids | 0.708 |  |  | 0.224 |  |  |  |  |  |
| cbcl_q07_p |  | Bragging, boasting | 0.443 |  |  | 0.302 |  |  |  |  |  |
| cbcl_q33_p |  | Feels or complains that no one loves them | 0.692 |  |  | 0.068 |  |  |  |  |  |
| cbcl_q19_p |  | Demands a lot of attention | 0.686 |  |  | 0.185 |  |  |  |  |  |
| cbcl_q74_p |  | Showing off or clowning | 0.536 |  |  | 0.246 |  |  |  |  |  |
| cbcl_q87_p |  | Sudden changes in mood or feelings | 0.805 |  |  | 0.059 |  |  |  |  |  |
| cbcl_q109_p |  | Whining | 0.553 |  |  | 0.155 |  |  |  |  |  |
| OftenArguesWithAdultsAuthority | ABCD Parent Diagnostic Interview for DSM-5 Full (KSADS-5) | Symptom - Often argues with adults/authority (Past or Present) | 0.576 |  |  | 0.629 |  |  |  |  |  |
| OftenLosesTemper |  | Symptom - Often loses temper  (Past or Present) | 0.558 |  |  | 0.395 |  |  |  |  |  |
| OftenDisobeysRulesRequests |  | Symptom - Often disobeys rules/requests  (Past or Present) | 0.559 |  |  | 0.524 |  |  |  |  |  |
| DurationAtLeast6MonthsODD |  | Duration (at least 6 months) ODD, Past | 0.584 |  |  | 0.523 |  |  |  |  |  |
| OftenBulliesOthers |  | Symptom - Often bullies others  (Past or Present) | 0.464 |  |  | 0.448 |  |  |  |  |  |
| Stealing |  | Symptom – Stealing (Past or Present) | 0.469 |  |  | 0.350 |  |  |  |  |  |
| OftenLies |  | Symptom - Often Lies (Past or Present) | 0.504 |  |  | 0.351 |  |  |  |  |  |
| cbcl_q111_p | ABCD Parent Child Behavior Checklist (CBCL)  ABCD Parent Child Behavior Checklist (CBCL) | Withdrawn, doesn't get involved with others | 0.583 |  |  |  | 0.514 |  |  |  |  |
| cbcl_q75_p |  | Too shy or timid | 0.301 |  |  |  | 0.612 |  |  |  |  |
| cbcl_q42_p |  | Would rather be alone than with others | 0.460 |  |  |  | 0.418 |  |  |  |  |
| cbcl_q71_p |  | Self-conscious or easily embarrassed | 0.475 |  |  |  | 0.556 |  |  |  |  |
| cbcl_q35_p |  | Feels worthless or inferior | 0.633 |  |  |  | 0.395 |  |  |  |  |
| cbcl_q50_p |  | Too fearful or anxious | 0.531 |  |  |  | 0.565 |  |  |  |  |
| cbcl_q65_p |  | Refuses to talk | 0.562 |  |  |  | 0.318 |  |  |  |  |
| cbcl_q103_p |  | Unhappy, sad, or depressed | 0.687 |  |  |  | 0.360 |  |  |  |  |
| cbcl_q112_p |  | Worries | 0.483 |  |  |  | 0.577 |  |  |  |  |
| cbcl_q52_p |  | Feels too guilty | 0.478 |  |  |  | 0.559 |  |  |  |  |
| cbcl_q102_p |  | Underactive, slow moving, or lacks energy | 0.550 |  |  |  | 0.304 |  |  |  |  |
| cbcl_q30_p |  | Fears going to school | 0.524 |  |  |  | 0.411 |  |  |  |  |
| cbcl_q32_p |  | Feels they have to be perfect | 0.281 |  |  |  | 0.531 |  |  |  |  |
| cbcl_q69_p |  | Secretive, keeps things to self | 0.582 |  |  |  | 0.279 |  |  |  |  |
| cbcl_q05_p |  | There is very little they enjoy | 0.653 |  |  |  | 0.237 |  |  |  |  |
| cbcl_q12_p |  | Complains of loneliness | 0.627 |  |  |  | 0.281 |  |  |  |  |
| cbcl_q31_p |  | Fears they might think or do something bad | 0.485 |  |  |  | 0.427 |  |  |  |  |
| cbcl_q45_p |  | Nervous, high strung, or tense | 0.606 |  |  |  | 0.438 |  |  |  |  |
| FearOfSocialSituations | ABCD Parent KSADS-5 | Symptom - Fear of Social Situations  (Past or Present) | 0.323 |  |  |  | 0.493 |  |  |  |  |
| cbcl_q08_p | ABCD Parent Child Behavior Checklist (CBCL) | Can't concentrate, can't pay attention for long | 0.682 |  |  |  |  | 0.632 |  |  |  |
| cbcl_q78_p |  | Inattentive or easily distracted | 0.694 |  |  |  |  | 0.585 |  |  |  |
| cbcl_q10_p |  | Can't sit still, restless, or hyperactive | 0.679 |  |  |  |  | 0.446 |  |  |  |
| cbcl_q04_p |  | Fails to finish things they start | 0.679 |  |  |  |  | 0.330 |  |  |  |
| cbcl_q61_p |  | Poor school work | 0.649 |  |  |  |  | 0.317 |  |  |  |
| cbcl_q17_p |  | Daydreams or gets lost in their thoughts | 0.523 |  |  |  |  | 0.308 |  |  |  |
| cbcl_q62_p |  | Poorly coordinated or clumsy | 0.574 |  |  |  |  | 0.195 |  |  |  |
| cbcl_q41_p |  | Impulsive or acts without thinking | 0.784 |  |  |  |  | 0.225 |  |  |  |
| cbcl_q80_p |  | Stares blankly | 0.622 |  |  |  |  | 0.283 |  |  |  |
| cbcl_q13_p |  | Confused or seems to be in a fog | 0.638 |  |  |  |  | 0.252 |  |  |  |
| cbcl_q01_p |  | Acts too young for their age | 0.561 |  |  |  |  | 0.231 |  |  |  |
| easily_distracted | ABCD Parent Diagnostic Interview for DSM-5 Full (KSADS-5) | Symptom - Easily distracted (Past or Present + for more than one school year) | 0.604 |  |  |  |  | 0.718 |  |  |  |
| diff_remain_seated |  | Symptom - Difficulty remaining seated (Past or Present + for more than one school year) | 0.610 |  |  |  |  | 0.684 |  |  |  |
| impulsivity |  | Symptom – Impulsivity (Past or Present + for more than one school year) | 0.673 |  |  |  |  | 0.531 |  |  |  |
| PoorEyeContact |  | Symptom - Poor Eye Contact (Past or Present) | 0.514 |  |  |  |  | 0.226 |  |  |  |
| UnusualBodyMovements |  | Symptom - Unusual body movements  (Past or Present) | 0.430 |  |  |  |  | 0.275 |  |  |  |
| cbcl_q56c_p | ABCD Parent Child Behavior Checklist (CBCL) | Nausea, feels sick | 0.423 |  |  |  |  |  | 0.800 |  |  |
| cbcl_q56f_p |  | Stomachaches | 0.373 |  |  |  |  |  | 0.730 |  |  |
| cbcl_q56g_p |  | Vomiting, throwing up | 0.310 |  |  |  |  |  | 0.624 |  |  |
| cbcl_q56b_p |  | Headaches | 0.340 |  |  |  |  |  | 0.518 |  |  |
| cbcl_q53_p | ABCD Parent CBCL | Overeating | 0.440 |  |  |  |  |  |  | 0.836 |  |
| cbcl_q55_p |  | Overweight | 0.247 |  |  |  |  |  |  | 0.836 |  |
| BingeEating | ABCD Parent KSADS-5 | Symptom - Binge Eating (Past or Present) | 0.457 |  |  |  |  |  |  | 0.628 |  |
| DistressAtInternalRemindersOfTrauma | ABCD Parent Diagnostic Interview for DSM-5 Full (KSADS-5) | Symptom - Distress at internal reminders of trauma (Past or Present) | 0.414 |  |  |  |  |  |  |  | 0.821 |
| EffortsToAvoidsThoughtsOfTrauma |  | Symptom - Efforts to avoids thoughts of trauma (Past or Present) | 0.402 |  |  |  |  |  |  |  | 0.789 |
| Nightmares |  | Symptom – Nightmares (Past or Present) | 0.481 |  |  |  |  |  |  |  | 0.650 |
| ExcessiveWorriesMoreDaysThanNot |  | Symptom - Excessive worries more days than not (Past or Present) | 0.530 |  |  |  |  |  |  |  | 0.339 |
| PanicAttacks |  | Symptom - Panic Attacks (Past or Present) | 0.476 |  |  |  |  |  |  |  | 0.301 |
| Insomnia |  | Symptom – Insomnia (Past or Present) | 0.499 |  |  |  |  |  |  |  | 0.192 |

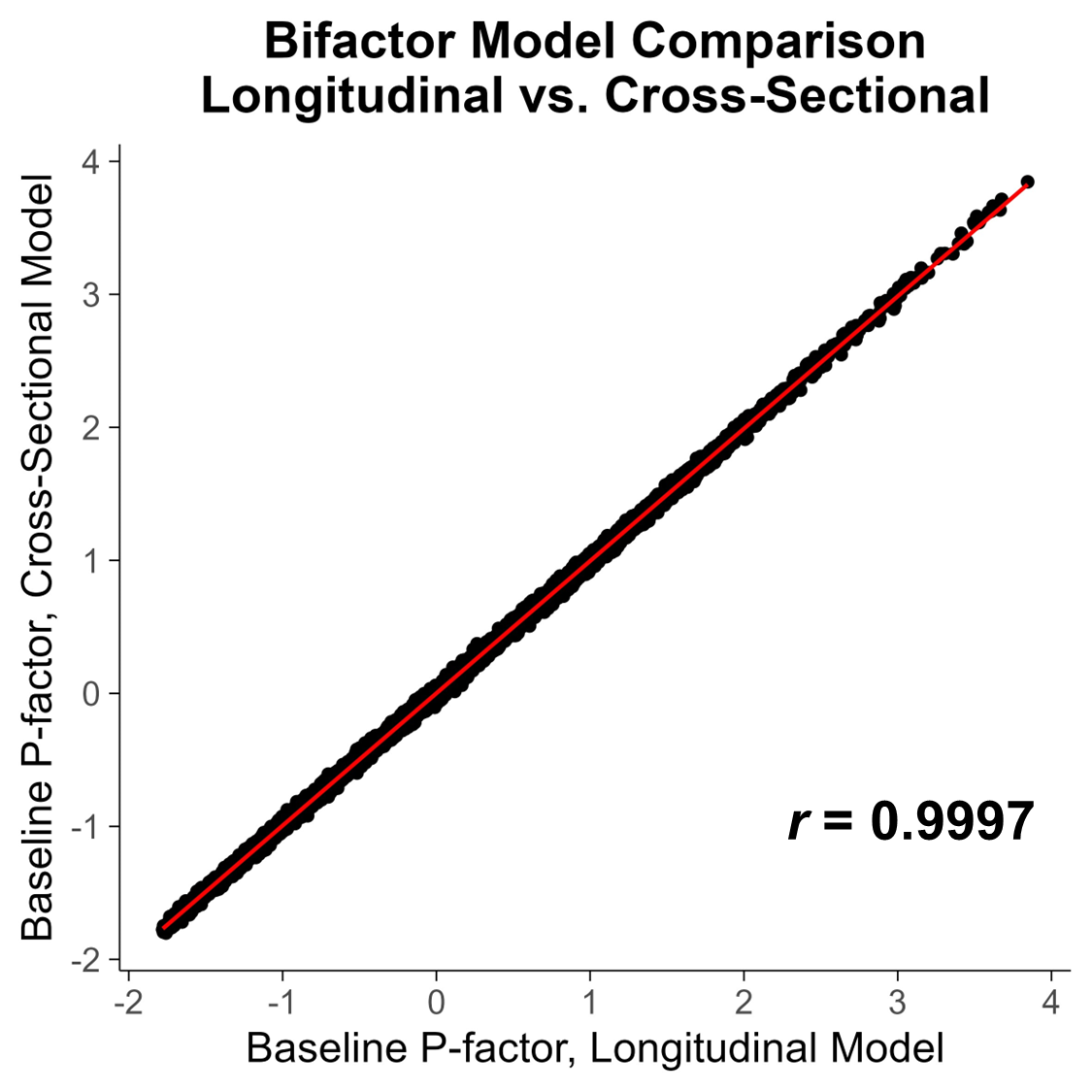

**eFig 1. Correlation between P-factor Scores from Longitudinal vs. Cross-sectional Bifactor Models.** Baseline p-factor derived from longitudinal bifactor analysis (including data from both baseline and year 2 time points) was highly correlated to baseline p-factor derived from a cross-sectional bifactor analysis (including only data from the baseline time point).

| **eTable 2. P-factor and PRS-F1 LME model** | | | | |
| --- | --- | --- | --- | --- |
| *Response* | **P-factor** | | | |
| *Predictors* | *Estimates* | *Std. Error* | *t* | *p* |
| Intercept | -0.12 | 0.02 | -6.64 | **3.32e-11** |
| PRS-F1 | 0.11 | 0.01 | 8.68 | **5.01e-18** |
| Age | 0.00 | 0.01 | 0.25 | 8.01e-01 |
| Biological Sex | 0.22 | 0.02 | 8.80 | **1.78e-18** |
| **Random Effects** | | | | |
| σ^2^ | 0.47 | | | |
| τ_00_ _rel_family_id_ | 0.41 | | | |
| ICC | 0.47 | | | |
| N _rel_family_id_ | 4819 | | | |
| Observations | 5815 | | | |
| Marginal R^2^ / Conditional R^2^ | 0.026 / 0.481 | | | |

| **eTable 3. P-factor and PRS-F2 LME model** | | | | |
| --- | --- | --- | --- | --- |
| *Response* | **P-factor** | | | |
| *Predictors* | *Estimates* | *Std. Error* | *t* | *p* |
| Intercept | -0.12 | 0.02 | -6.58 | **4.98e-11** |
| PRS-F2 | 0.02 | 0.01 | 1.80 | 7.23e-02 |
| Age | 0.00 | 0.01 | 0.34 | 7.34e-01 |
| Biological Sex | 0.22 | 0.02 | 8.78 | **2.14e-18** |
| **Random Effects** | | | | |
| σ^2^ | 0.47 | | | |
| τ_00_ _rel_family_id_ | 0.42 | | | |
| ICC | 0.47 | | | |
| N _rel_family_id_ | 4819 | | | |
| Observations | 5815 | | | |
| Marginal R^2^ / Conditional R^2^ | 0.014 / 0.481 | | | |

| **eTable 4. PRS-F1 and PRS-F2 LME model** | | | | |
| --- | --- | --- | --- | --- |
| Response | **PRS-F2** | | | |
| *Predictors* | *Estimates* | *Std. Error* | *t* | *p* |
| Intercept | 0.01 | 0.02 | 0.31 | 7.55e-01 |
| PRS-F1 | 0.18 | 0.01 | 14.70 | **4.41e-48** |
| Age | 0.00 | 0.01 | 0.15 | 8.82e-01 |
| Biological Sex | -0.02 | 0.02 | -0.70 | 4.81e-01 |
| **Random Effects** | | | | |
| σ^2^ | 0.31 | | | |
| τ_00_ _rel_family_id_ | 0.49 | | | |
| ICC | 0.61 | | | |
| N _rel_family_id_ | 4819 | | | |
| Observations | 5815 | | | |
| Marginal R^2^ / Conditional R^2^ | 0.036 / 0.625 | | | |

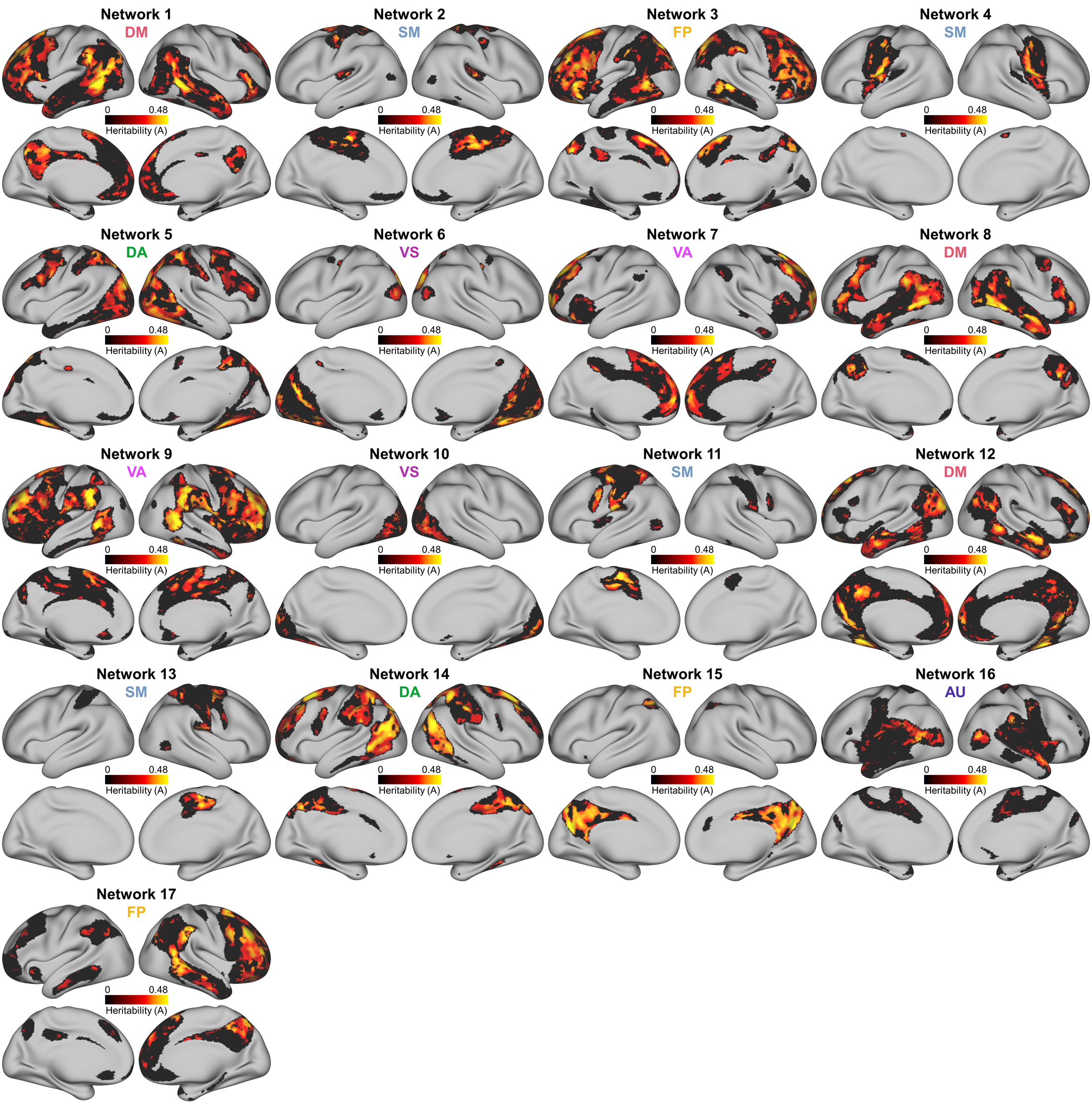

**eFig 2. Significant Heritabilities of PFN vertex loadings across 17 networks.** Individual maps showing significant heritability of PFN vertex network loadings, defined as the heritability of a specific vertex belonging to a specific network, thresholded by significance with false discovery rate (FDR) correction (*p*<0.05). Of the 235,355 non-zero variance vertex loadings, 79,835 (33.9%) are significantly heritable.

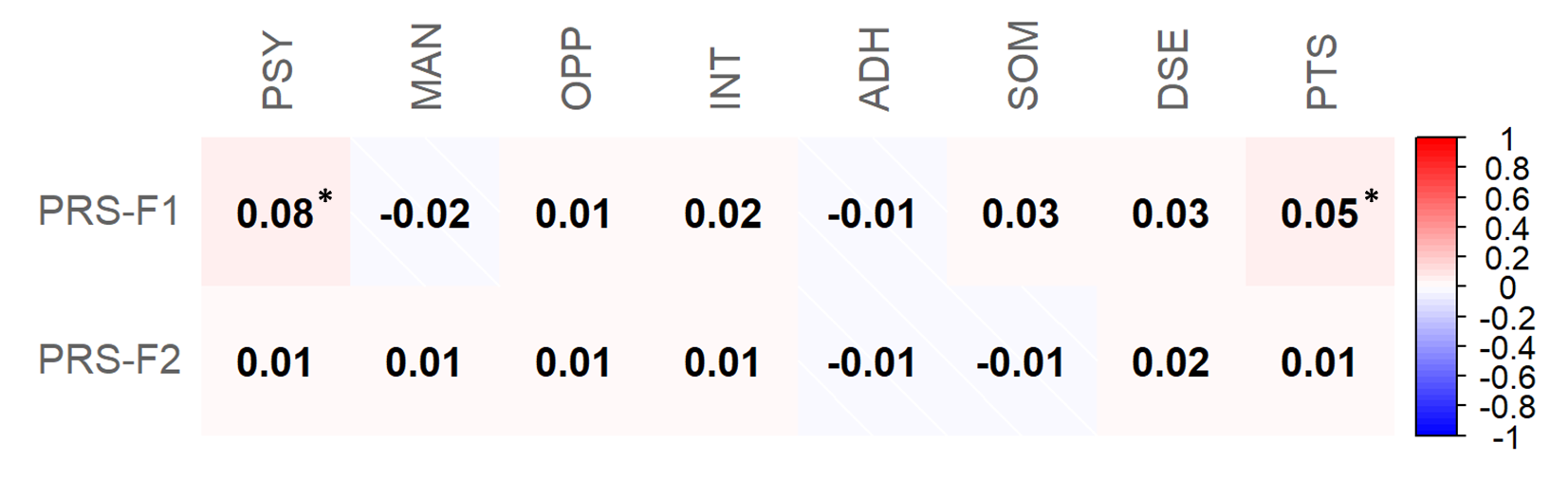
**eFig 3. PRS-F1 and PRS-F2 Correlations with the 8 Sub-factor Scores**. To test if polygenic risk of F1 or F2 was related to symptom variability beyond that captured by p-factor, we calculated Pearson correlations of PRS-F1 and PRS-F2 with sub-factor scores orthogonal to p-factor (PSY = psychotic symptoms, MAN = manic symptoms, OPP = oppositional defiance, INT = internalizing symptoms, ADH = attention-deficiency and hyperactivity, SOM = somatic symptoms, DSE = disordered eating, PTS = post-traumatic stress). Significant correlations were found between PRS-F1 and the PSY and PTS sub-factors; no significant correlations were found between PRS-F2 and any sub-factors. ****p<0.05 after false discovery rate (FDR) correction.***

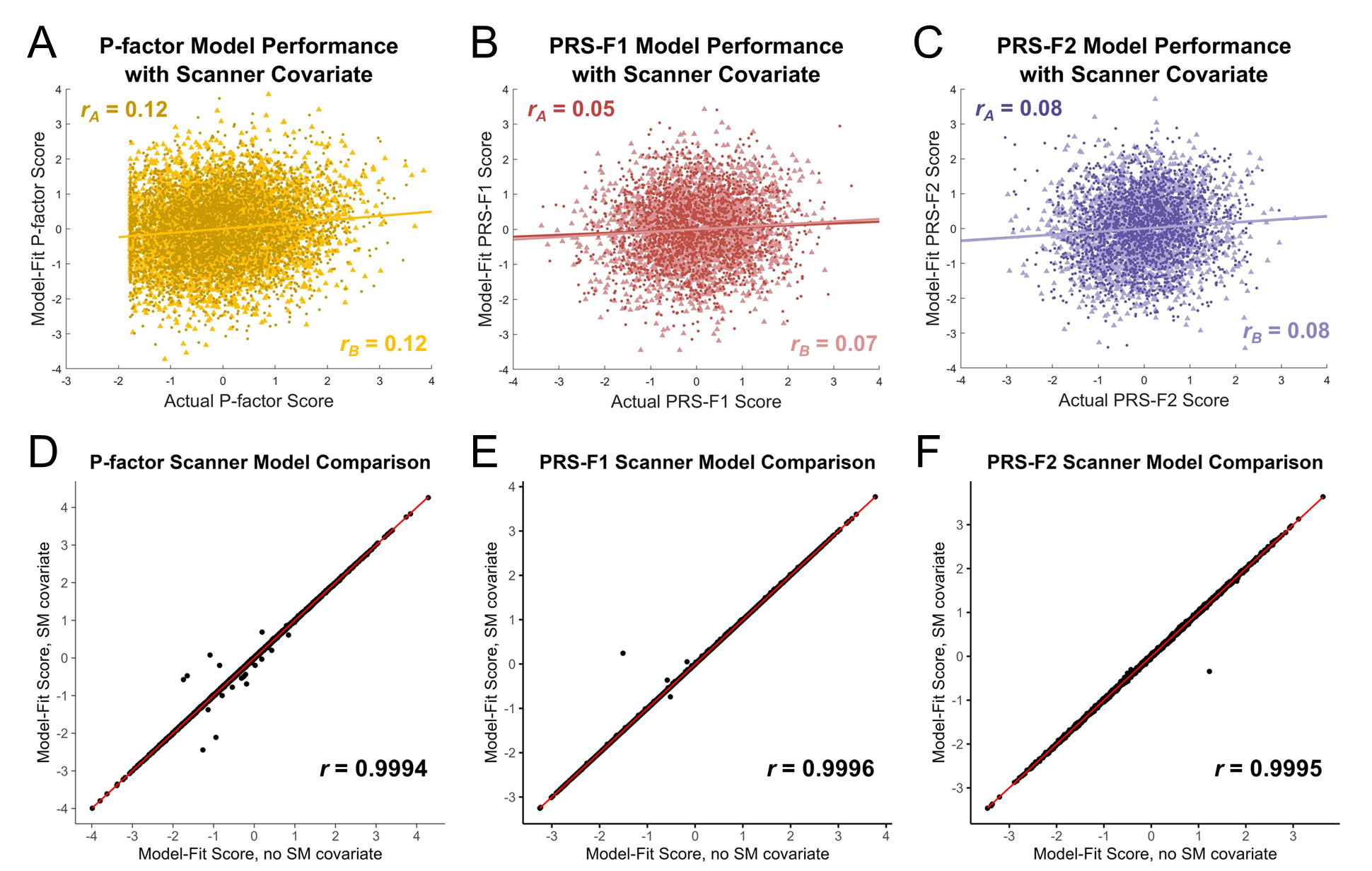

**eFig 4. Ridge Regression Models with inclusion of Scanner Manufacturer as a Covariate.** When scanner manufacturer was included as a covariate in ridge regression models, multivariate associations between PFN topography and p-factor (**A**), PRS-F1 (**B**), and PRS-F2 (**C**), as defined by model performance, were not affected compared to original models without inclusion of the scanner covariate (**Fig 3A-C**). Model-fit scores from ridge regression models including scanner manufacturer (SM) covariate were highly correlated to model-fit scores from original models without SM covariate in the case of p-factor (**D**), PRS-F1 (**E**), and PRS-F2 (**F**).

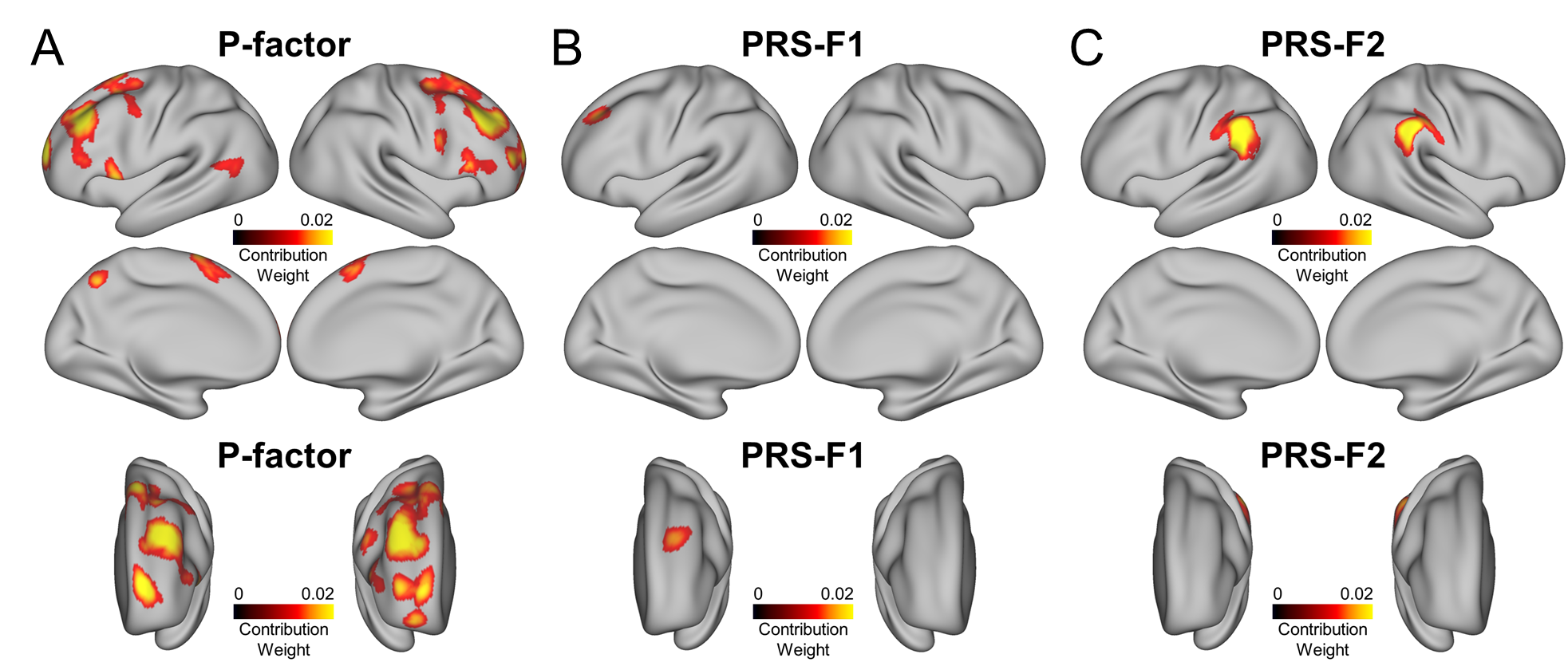

**eFig 5. Significance Testing of Cortical Regions Driving PFN Associations with P-factor, PRS-F1, and PRS-F2.** At each vertex, absolute Haufe-transformed contribution weights were summed across the 17 networks and the resulting sums were processed using threshold-free cluster enhancement (TFCE), an approach which integrates signal cluster height and spatial extent over all thresholds^41^. The TFCE scores were compared against a null distribution of maximal TFCE values derived from 1000 participant-level permutations. Here, we show the contribution weights of vertices that survived family-wise error correction (*p_FWE_*<0.05) for PFN models of p-factor (**A**), PRS-F1 (**B**), and PRS-F2 (**C**), corresponding to un-thresholded maps shown in **Fig 4A-C**. Bottom row shows prefrontal cortex view.

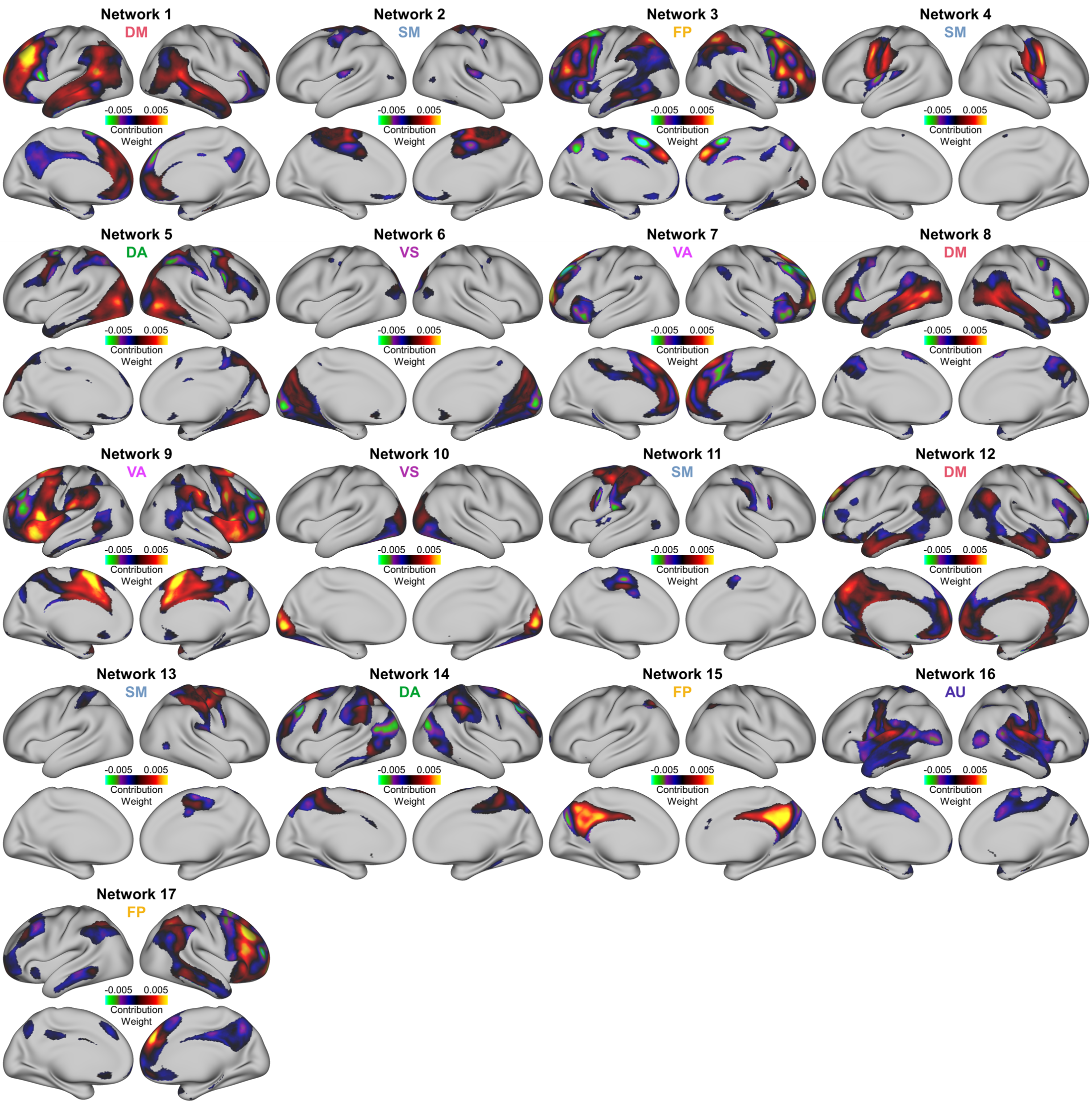
**eFig 6. P-factor Weight Maps**. Cortical maps showing the Haufe-transformed contribution weights of each network (each corresponds to one of 17 columns in the PFN loading matrix) for the p-factor ridge regression model. (FP = frontoparietal, VA = ventral attention, DA = dorsal attention, DM = default mode, AU = auditory, SM = somatomotor, VS = visual).

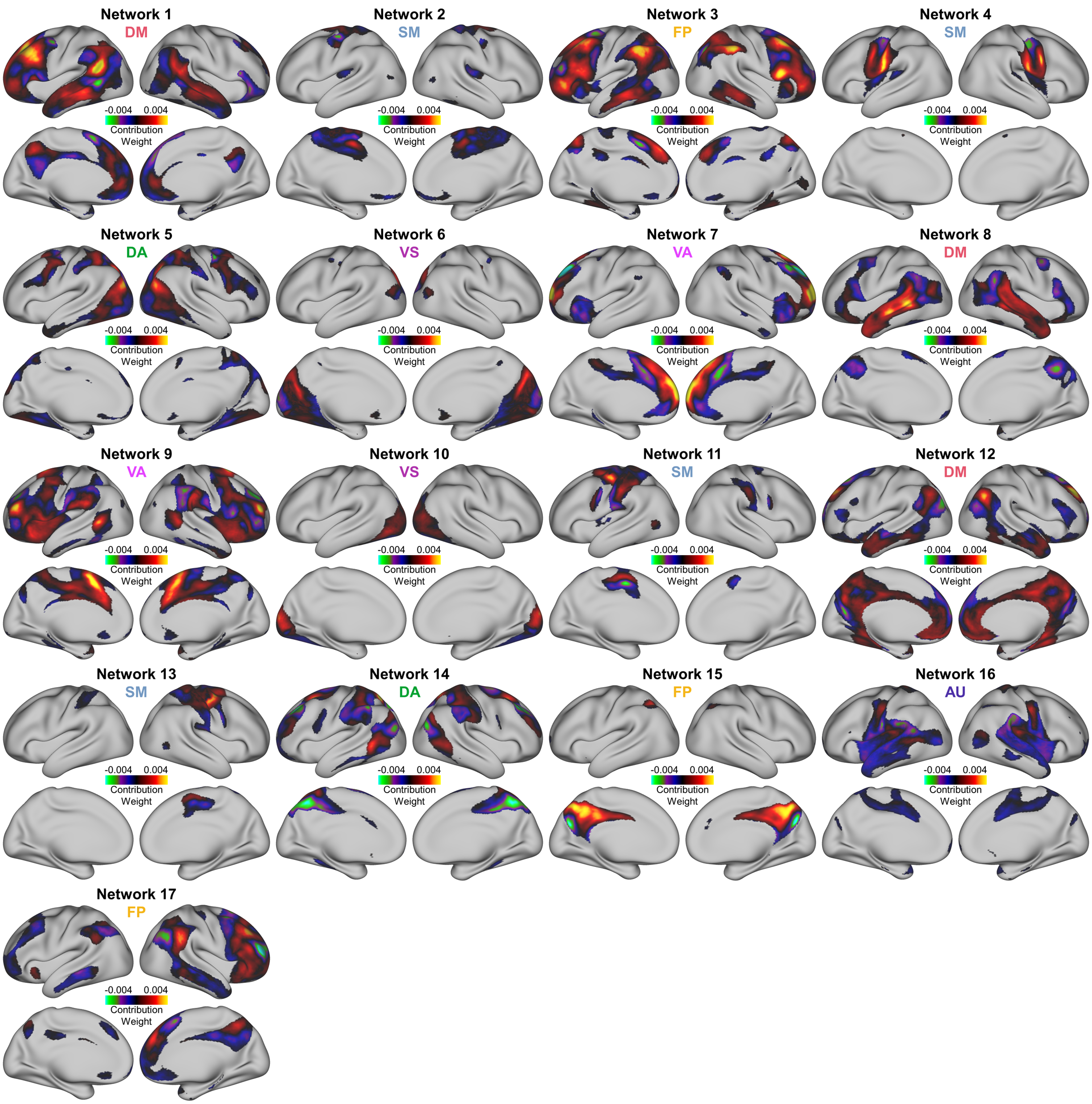
**eFig 7. PRS-F1 Weight Maps**. Cortical maps showing the Haufe-transformed contribution weights of each network (each corresponds to one of 17 columns in the PFN loading matrix) for the PRS-F1 ridge regression model. (FP = frontoparietal, VA = ventral attention, DA = dorsal attention, DM = default mode, AU = auditory, SM = somatomotor, VS = visual).

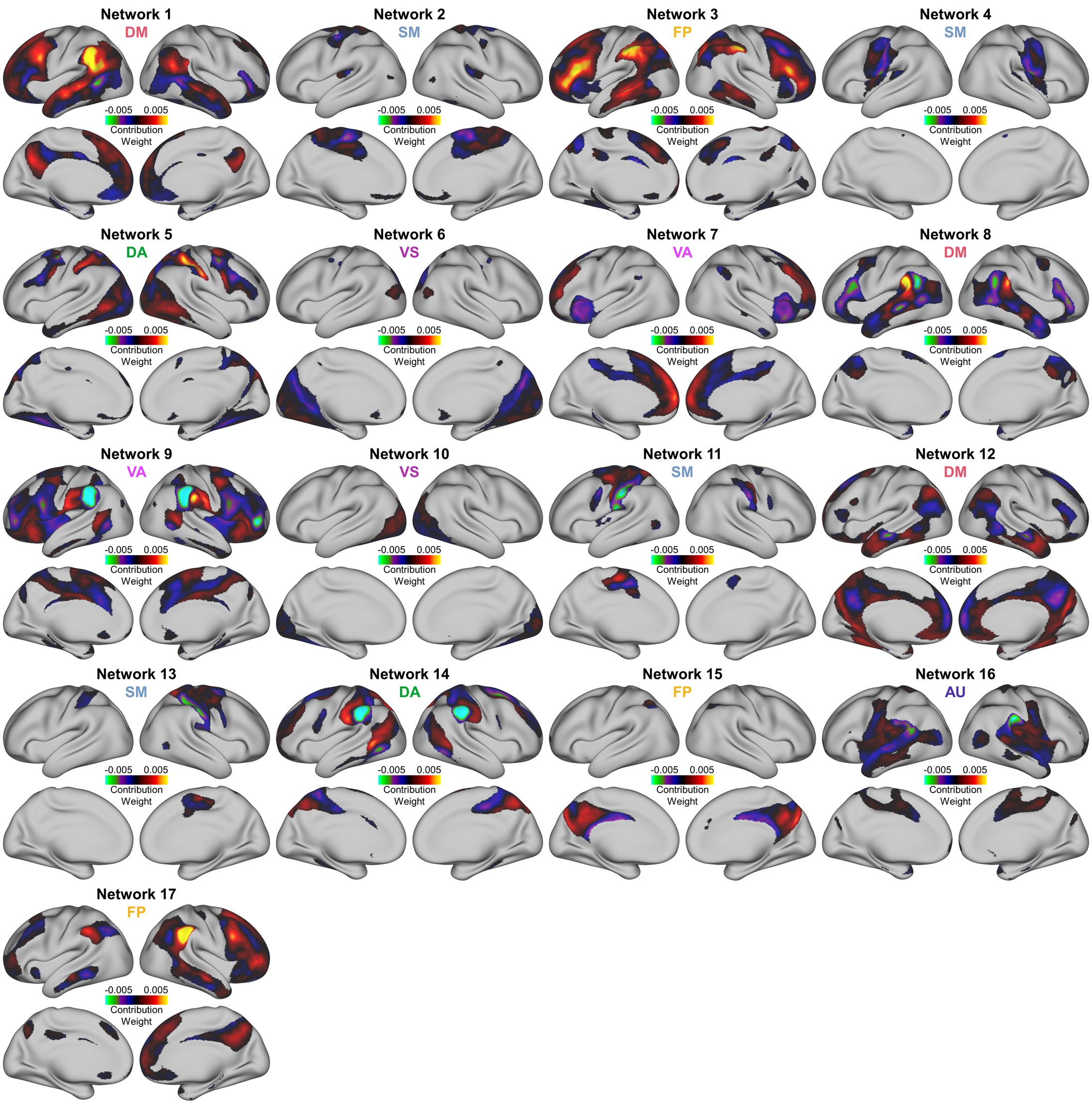

**eFig 8. PRS-F2 Weight Maps**. Cortical maps showing the Haufe-transformed contribution weights of each network (each corresponds to one of 17 columns in the PFN loading matrix) for the PRS-F2 ridge regression model. (FP = frontoparietal, VA = ventral attention, DA = dorsal attention, DM = default mode, AU = auditory, SM = somatomotor, VS = visual).

**eFig 9. Top 5% and 1% Directional Network Topograph**
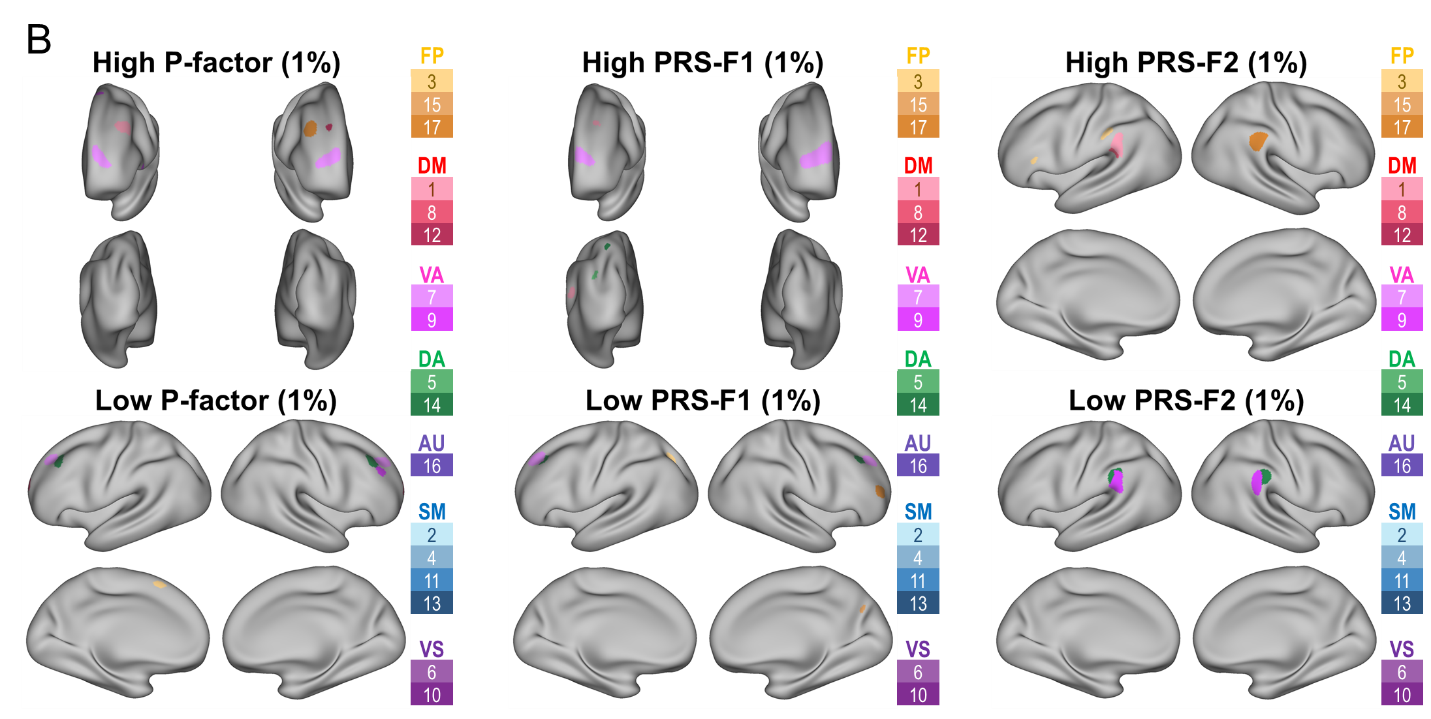

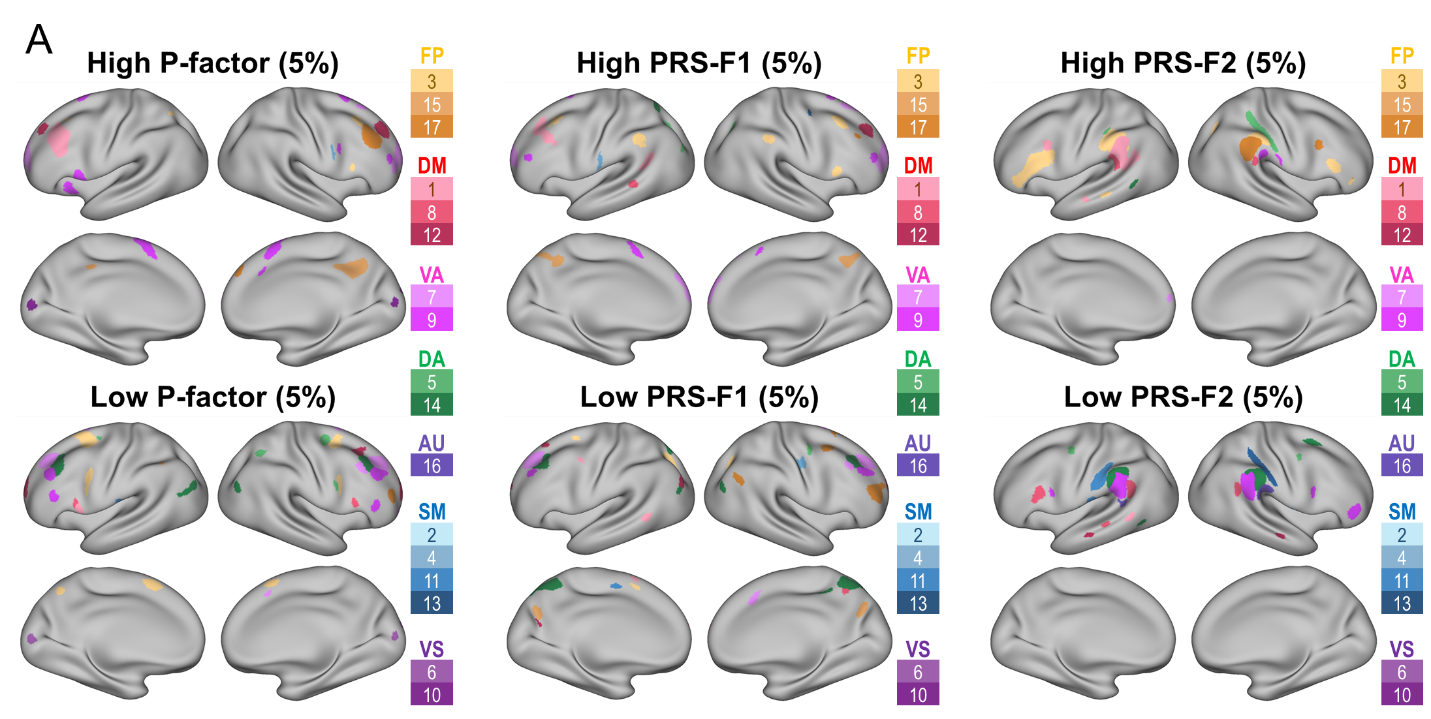
**y Maps**. **A top.** To identify network topography positively associated with our variables of interest, we determined the most positive Haufe-transformed model weight across the 17 networks for each vertex, thresholded to keep the top 5% of most positive weights across the cortical surface, and set a 25mm^2^ cluster threshold. Then, we assigned each of the retained vertices to the network corresponding to each vertex’s most positive weight (network denoted by specific color as shown in legend on the right of each panel). **A bottom.** To identify network topography negatively associated with our variables of interest, we determined the most negative Haufe-transformed model weight across the 17 networks for each vertex, thresholded to keep the top 5% of values (by magnitude) across the cortical surface, and set a 25mm^2^ cluster threshold. Then, we assigned each of the retained vertices to the network corresponding to each vertex’s most negative weight (network denoted by specific color as shown in legend on the right of each panel). **B.** Same as A, except thresholded to keep the top 1% of most positive (**top**) and most negative weights (**bottom**).

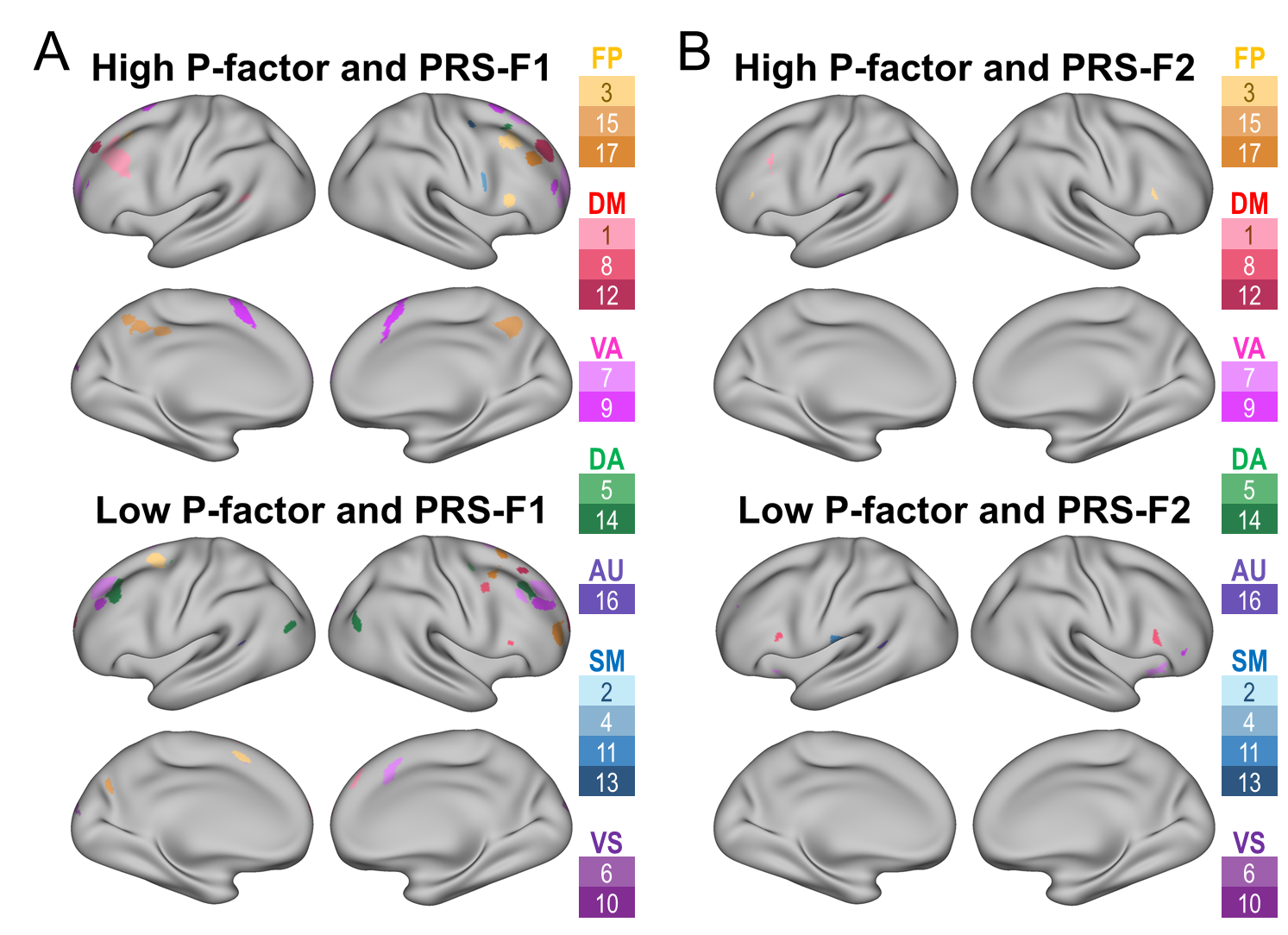

**eFig 10. Conjunction Maps of Top 10% Directional Network Topographies. A.** Conjunction of top 10% directional topography maps of p-factor (Fig 5A middle, bottom) and PRS-F1 (Fig 5B middle, bottom) **Top.** High P-factor and PRS-F1 share 2,456/5,941 vertices (41.3%) of their top 10% network identities (whole cortex view of Fig 5E). **Bottom.** Low P-factor and PRS-F1 share 1,711/5,941 vertices (28.8%) of their top 10% network identities (whole cortex view of Fig 5F). **B.** Conjunction of top 10% directional topography maps of p-factor (Fig 5A middle, bottom) and PRS-F2 (Fig 5C middle, bottom). **Top.** High P-factor and PRS-F2 share only 125/5,941 vertices (2.1%) of their top 10% network identities. **Bottom.** Low P-factor and PRS-F2 share only 269/5,941 vertices (4.5%) of their top 10% network identities.
